# A Novel Digital Twin Strategy to Examine the Implications of Randomized Clinical Trials for Real-World Populations

**DOI:** 10.1101/2024.03.25.24304868

**Authors:** Phyllis M. Thangaraj, Sumukh Vasisht Shankar, Sicong Huang, Girish N. Nadkarni, Bobak J. Mortazavi, Evangelos K. Oikonomou, Rohan Khera

## Abstract

Randomized clinical trials (RCTs) are essential to guide medical practice; however, their generalizability to a given population is often uncertain. We developed a statistically informed Generative Adversarial Network (GAN) model, RCT-Twin-GAN, that leverages relationships between covariates and outcomes and generates a digital twin of an RCT (RCT-Twin) conditioned on covariate distributions from a second patient population. We used RCT-Twin-GAN to reproduce treatment effect outcomes of the Systolic Blood Pressure Intervention Trial (SPRINT) and the Action to Control Cardiovascular Risk in Diabetes (ACCORD) Blood Pressure Trial, which tested the same intervention but found different treatment effects. To demonstrate treatment effect estimates of each RCT conditioned on the other RCT’s patient population, we evaluated the cardiovascular event-free survival of SPRINT digital twins conditioned on the ACCORD cohort and vice versa (ACCORD twins conditioned on SPRINT). The conditioned digital twins were balanced across intervention and control arms (mean absolute standardized mean difference (MASMD) of covariates between treatment arms 0.019 (SD 0.018), and the conditioned covariates of the SPRINT-Twin on ACCORD were more similar to ACCORD than SPRINT (MASMD 0.0082 SD 0.016 vs. 0.46 SD 0.20). Notably, across iterations, SPRINT conditioned ACCORD-Twin datasets reproduced the overall non-significant effect size seen in ACCORD (5-year cardiovascular outcome hazard ratio (95% confidence interval) of 0.88 (0.73-1.06) in ACCORD vs. median 0.87 (0.68-1.13) in the SPRINT conditioned ACCORD-Twin), while the ACCORD conditioned SPRINT-Twins reproduced the significant effect size seen in SPRINT (0.75 (0.64-0.89) vs. median 0.79 (0.72-0.86)) in the ACCORD conditioned SPRINT-Twin). Finally, we demonstrate the translation of this approach to real-world populations by conditioning the trials on an electronic health record population. Therefore, RCT-Twin-GAN simulates the direct translation of RCT-derived treatment effects across various patient populations.

## INTRODUCTION

Randomized clinical trials (RCTs) generate evidence that defines optimal clinical practices, but their generalizability to real-world patient populations is often challenging to quantify.^1,2^ This is a concern because RCTs often have underrepresentation from several demographic and clinical subpopulations^3–7^ and varying treatment effects among individuals with certain characteristics.^8–10^ These considerations are critical to translating information from RCTs to real-world patient populations,^11,12^ but no strategies exist to evaluate how they may affect the applicability to patients in these settings.

Variation across RCTs testing similar interventions with discrepant treatment effects is a key issue for the generalizability of interventions tested in RCTs.^13–19^ For example, the Systolic Blood Pressure Intervention Trial (SPRINT) was a treatment intervention RCT that showed improved cardiovascular outcomes with intensive blood pressure control.^13^ In contrast, the Action to Control Cardiovascular Risk in Diabetes Blood Pressure (ACCORD) trial did not find improved cardiovascular outcomes with the same intervention.^14^ Among the explanations posited for these discrepant findings include differences in population composition and event rates.^20–23^ Despite experimental evidence from two trials, there is no quantitative strategy to evaluate these assertions explicitly. Therefore, while it is critical to evaluate whether the effects observed in an RCT population generalize to a second population – either a planned second RCT or a general population of patients with the condition – the challenge remains to examine these effects in the context of the complex differences across multiple population characteristics.

Digital twins of RCTs introduce a strategy to create a synthetic representation of a clinical trial updated by attributes of a second population. Specifically, trial-level digital twin synthesis through deep generative models such as Generative Adversarial Networks (GANs) can integrate multiple covariates from a patient cohort by constructing a digital twin with covariate values sampled from the second cohort while retaining relationships and correlations between variables within the original RCT. While GANs have been utilized to estimate individual treatment effects, their potential for evidence translation across patient populations has not been explored.^24–27^ Conditional GANs (CGAN) enable the generation of synthetic datasets that condition a model with covariates from a second population distribution.^28,29^ We hypothesize that applying this model to an RCT conditioned on a second population will estimate the treatment effects of the original RCT in the new patient population.

We present RCT-Twin-GAN, a generative framework that combines clinical knowledge and the statistically informed architecture to create a digital twin of an RCT conditioned on the characteristics of a second patient population to assess for the generalizability of the treatment effect (Figure 1, Figure 2). To demonstrate the ability of the digital twin to replicate treatment effects in the conditioning population, we first compared two RCTs, SPRINT and ACCORD, with similar interventions but disparate treatment effects on cardiovascular outcomes. We created a digital twin of each of the 2 RCTs conditioned on covariate distributions of the other and evaluated whether the RCT-Twins reproduced the treatment effect of the conditioning cohort. Finally, we describe the cardiovascular outcomes of SPRINT and ACCORD digital twins conditioned on characteristics of patients in the electronic health record (EHR), introducing the role of RCT-Twins in estimating RCT treatment effects in real-world populations.

**Figure 1:**
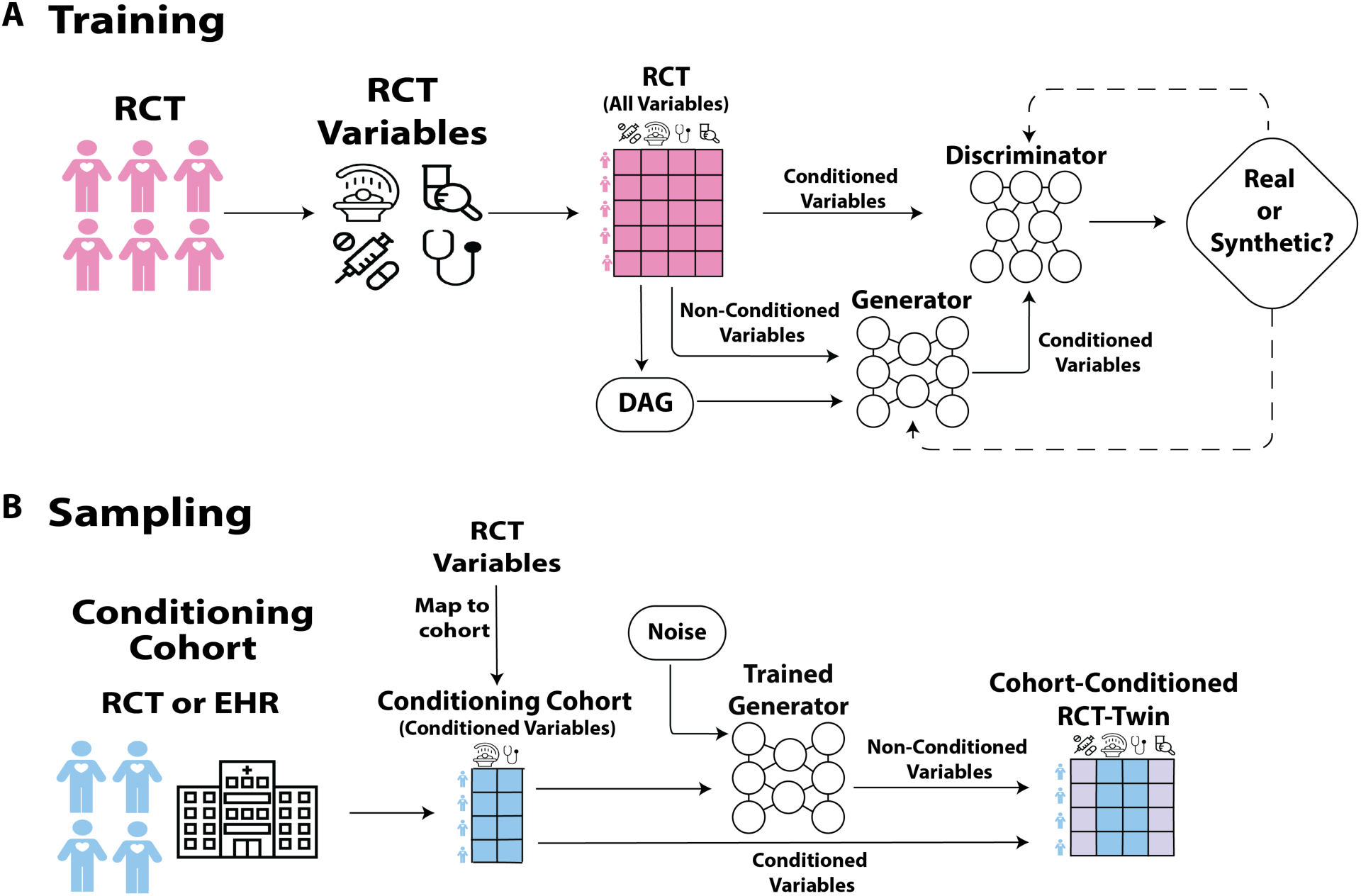
Graphical abstract of RCT-Twin-GAN model. A. In the training phase, the original cohort (pink) is a randomized clinical trial (RCT), and variables across all clinical domains are extracted from the cohort. The directed acyclic graph (DAG) includes clinician-defined relationships between original cohort covariates and is inputed to the generator, along with RCT values of the non-conditioned variables. The generator then creates the conditioned variables, and the discriminator must differentiate from the original RCT conditioned variables and the generator conditioned variables. Once the disciminator cannot distinguish between the original and generated values, the training is complete. B. In the sampling phase, conditioned variables from the RCT cohort are mapped to a conditioning cohort (blue), examples of which are another RCT or a patient cohort in the electronic health record (EHR). The trained generator then takes the conditioned variables from the conditioning cohort and noise as input, and then generates non-conditioned variables. The final cohort-conditioned RCT twin has conditioned covariate values from the conditioning cohort (blue) and generated non-conditioned covariates based off the relationships and correlations between covariates (light purple). Abbreviations: DAG: Directed Acyclic Graph, EHR: electronic health record, and RCT: Randomized Clinical Trial.

**Figure 2:**
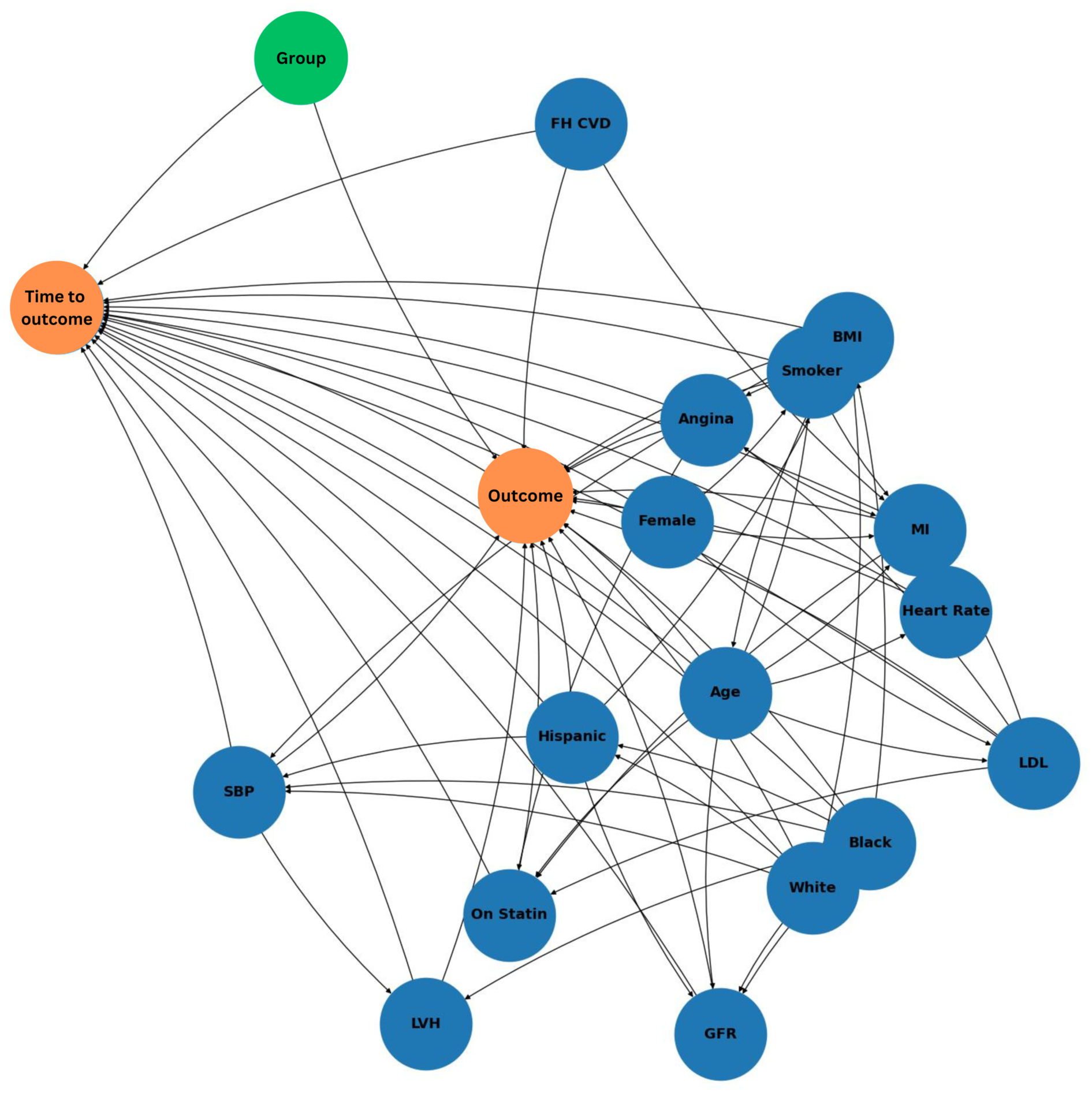
Directed acyclic graph of RCT-Twin-GAN. Directed relationships between covariates (in blue), time to outcome and outcome (in orange), and treatment arm designation (“Group”) in green. Abbreviations: BMI: Body Mass Index, CVD: Cardiovascular disease, FH: Family History, GFR: Glomerular Filtration Rate, LVH: Left ventricular hypertrophy, LDL: low-density lipoprotein, MI: Myocardial infarction, SBP: Systolic Blood Pressure.

## RESULTS

### Study Populations

The study developed digital twins of two RCTs. The first RCT, SPRINT, was a treatment intervention study to test whether intensive blood pressure control (goal systolic blood pressure less than 120 mmHg) versus standard care (goal systolic blood pressure less than 140 mmHg) reduced major cardiovascular events. The trial consisted of 9361 participants (median age 67 (61 to 76 (25-75% IQR, and 3332 (36%) women). The patients in SPRINT were followed for a median of 3.26 years for the first occurrence of any of the primary composite outcome of myocardial infarction, acute coronary syndrome, stroke, heart failure, or death from cardiovascular cause.

Our study built a SPRINT digital twin with a population representation of another RCT with the same intervention, the ACCORD trial, a double factorial RCT of participants with type 2 diabetes mellitus and cardiovascular disease. We specifically leveraged the blood pressure management component of the ACCORD trial, wherein half of the participants were randomized to intensive versus standard care blood pressure control, with the same treatment goals as those in the SPRINT trial. ACCORD consisted of 4733 participants (median age 62, IQR, 58-67, and 2258 [48%] women). ACCORD median follow-up time was 4.7 years for the primary composite outcome of myocardial infarction, stroke, or death from cardiovascular cause.

We also incorporated two cohorts from the Yale New Haven Hospital Health System Electronic Health Record (EHR), a large healthcare system including several hospitals with diverse racial and socioeconomic demographics across Connecticut and Rhode Island. Two sets of patients with hypertension, one without (N=22,132) and the other with diabetes (N=8,840) were identified to broadly represent populations included in SPRINT and ACCORD, respectively, to estimate the treatment effects found in the two RCTs on corresponding real-world patient populations. The final cohorts included 3,130 patients in the SPRINT EHR cohort and 2,731 patients in the ACCORD EHR cohort. The SPRINT EHR cohort had a median age of 73 years (IQR, 61 to 84) and 2069 (52%) women), while the ACCORD EHR cohort had a median age of 71 (IQR, 61 to 80) and 2032 (51%) women).

### The Non-Conditioned SPRINT Digital Twin Cohort

We created 10 SPRINT-Twins (the non-conditioned SPRINT twin), which had a median age of 66 (IQR, 60 to 75) and 1516-1704 (32-38%) women (Table S1, S2). The SPRINT-Twin reproduced the distributions of the original variables (covariates, outcome, and time to outcome) in SPRINT as evidenced by an absolute standardized mean difference (ASMD) of less than 0.1 for each variable and a mean absolute standardized difference (MASMD) of 0.020 (SD 0.015) between the SPRINT Control (C) Arm and SPRINT-Twins C Arm and 0.021 (SD 0.014) between the SPRINT Intervention (I) Arm and SPRINT-Twins I Arm. In addition, all variables were balanced between the I and C arms in the SPRINT-Twin, as evidenced by an ASMD of less than 0.1 for each variable and a MASMD of 0.011 (SD 0.016) between treatment arms across all variables. This was similar to the MASMD between treatment arms of SPRINT, 0.021 (SD 0.018) and below the threshold where distributions are considered substantially dissimilar. The correlations between variables were also preserved as evidenced by 88.4% concordance between the Spearman correlations calculated between SPRINT’s variables and those calculated between the SPRINT twin’s variables (Table S3, Figure S1).

### The Conditioned SPRINT_ACCORD_ and ACCORD_SPRINT_ Digital Twin Cohorts

We then generated 10 SPRINT_ACCORD_ Twins, which were SPRINT twins conditioned with values from the ACCORD cohort for 10 covariates, and 10 ACCORD_SPRINT_ Twins, which were ACCORD twins conditioned with values from the SPRINT cohort for 10 covariates. The SPRINT_ACCORD_ Twins had a median age of 62 years (IQR 58 to 68), 1106-1178 (46-49%) women (Tables S4, S5) with mean 2345 (SD 25.9) or 49.5% in the C arm, 2388 (SD 24) or 50.5% in the I arm. ACCORD_SPRINT_ Twins had a median age of 67 years (IQR 61 to 76), 1545-1677 (32-34%) women with mean 4759 (SD 66) or 50.8% in the C arm and 4603 (SD 62) or 49.2% in the I arm (Tables S6, S7). Across all treatment arm covariate distributions of the SPRINT_ACCORD_ Twins and ACCORD_SPRINT_ Twins, there was little difference between the I and C arms, suggesting balanced treatment arms as evidenced by each covariate having an ASMD between treatment arms of less than 0.1 with an MASMD between treatment arms of 0.024 (SD 0.017) for SPRINT_ACCORD_ Twins and 0.018 (SD 0.004) for ACCORD_SPRINT_ Twins, respectively (Figure 3a).

**Figure 3:**
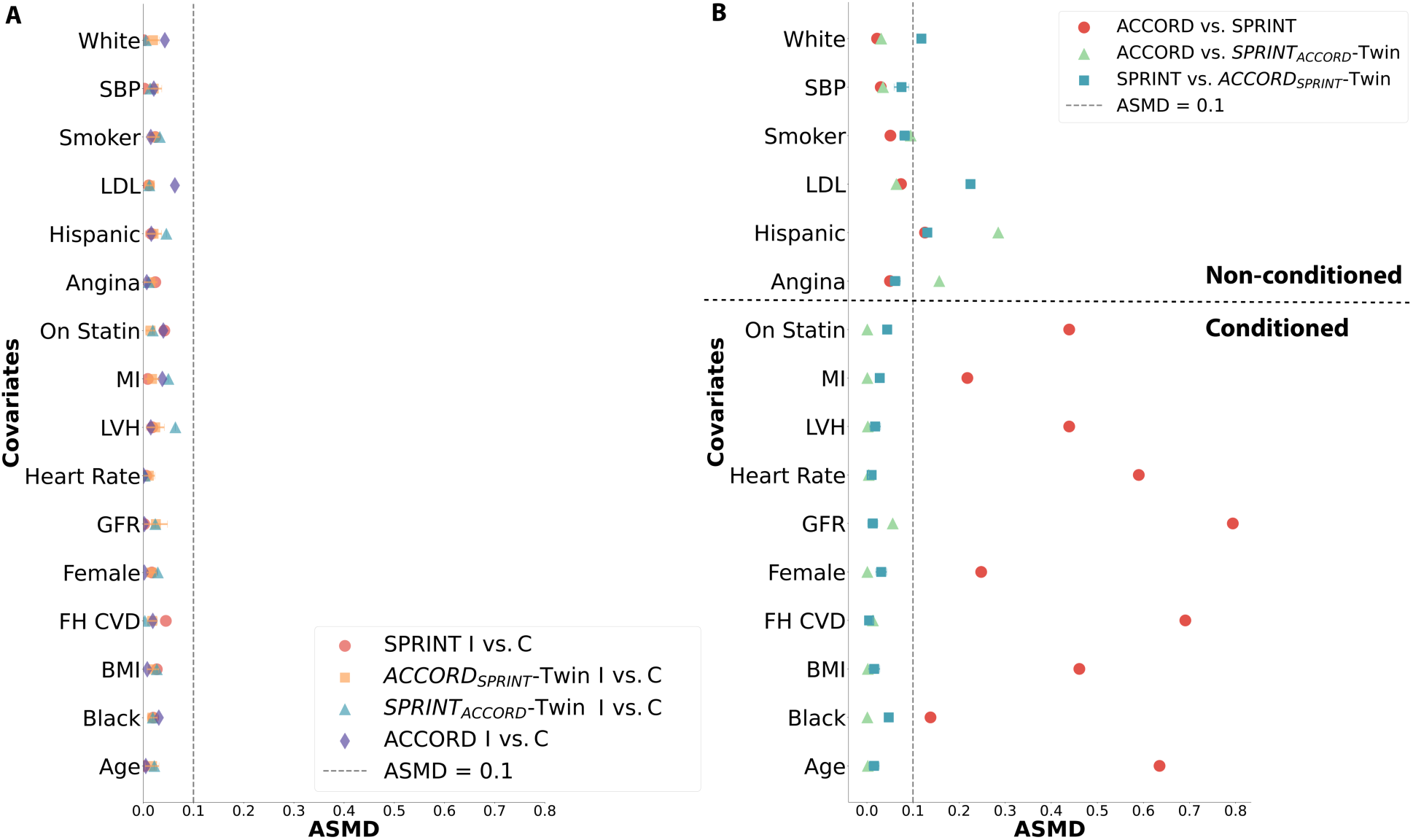
Absolute standardized mean difference (ASMD) of covariates between datasets. (A) ASMD of covariates between treatment arms of RCTs and digital twins. Markers include SPRINT (red circle), ACCORD_SPRINT_ Twin (orange square), SPRINT_ACCORD_-Twin (blue triangle), and ACCORD (purple diamond). The digital twin ASMDs are the mean of the 10 digital twin samples with standard deviation error bars. (B) ASMD of covariates between RCTs and digital twins. Red circle represents ASMD between ACCORD and SPRINT, green triangle represents ASMD between ACCORD and SPRINT_ACCORD_-Twins, and the blue square represents the ASMD between SPRINT and ACCORD_SPRINT_ Twins. The digital twin ASMDs are the mean of the 10 digital twin samples with standard deviation error bars. The grey dotted line represents an ASMD of 0.1, and the black dotted line separates non-conditioned and conditioned covariates. The conditioning covariates included Age, Black, BMI, FH CVD, Female, GFR, Heart Rate, LVH, MI, and On Statin. Abbreviations: ASMD: Absolute Standardized Mean Difference, BMI: Body Mass Index, CVD: Cardiovascular disease, C: Control Arm, FH: Family History, eGFR: Glomerular Filtration Rate, I: Intervention Arm, LDL: low-density lipoprotein, LVH: Left ventricular hypertrophy, MI: Myocardial infarction, SBP: Systolic Blood Pressure.

Comparing datasets, across all the conditioned covariates, the ASMD between the SPRINT_ACCORD_-Twin and ACCORD were less than 0.1, with a MASMD of 0.008 (SD 0.016), and similarly, the ASMDs between ACCORD_SPRINT_ Twin and SPRINT were less than 0.1 with a MASMD 0.023 (SD 0.014) compared to an MASMD of 0.46 (SD 0.20) for the same covariates between SPRINT and ACCORD (Figure 3b). Out of the six non-conditioned covariates, white race, systolic blood pressure, smoker, and LDL cholesterol level had ASMDs less than 0.1 between ACCORD vs. SPRINT_ACCORD_ Twin while systolic blood pressure, smoker, and angina had ASMDs less than 0.1 between SPRINT and ACCORD_SPRINT_ Twin (Figure 3b). Conversely, when conditioning on the opposite RCT and comparing datasets (ie. ACCORD vs ACCORD_SPRINT_ Twin and SPRINT vs SPRINT_ACCORD_ Twin), the ASMD resembles ACCORD vs SPRINT for the conditioned covariates (Figure S2). Similar to the non-conditioned twins, the correlations between variables were also preserved, as evidenced by the 85.6% concordance of the Spearman correlations between the ACCORD variables and the correlations between the SPRINT_ACCORD_ Twin variables, the 78.4% concordance of correlations between the SPRINT variables and the correlations between the SPRINT_ACCORD_ Twin variables, the 84.5% concordance of the correlations between the ACCORD variables and the ACCORD_SPRINT_ twin variables, and the 85.6% concordance of the correlations between the SPRINT variables and the correlations between the ACCORD_SPRINT_ Twin variables (Table S3, Figure S1).

### Digital Twin Similarity Evaluation

Given the generated nature of the complementary covariates, each row of the conditioned twins does not perfectly match the original cohort patients since it is updated with the conditioning cohort data, so covariate distribution level assessments were conducted. When training a multivariate logistic regression classifier to distinguish between RCT and twin data, in which an accuracy of 0.5 is considered random chance, we found the median accuracy of the model to correctly classify the data as real or fake to be 0.50 (IQR 0.49 to 0.51) for distinguishing SPRINT from SPRINT Twins, 0.50 (IQR 0.49 to 0.51) for distinguishing SPRINT from SPRINT_ACCORD_ Twins, 0.50 (IQR 0.49 to 0.55) for distinguishing ACCORD from ACCORD Twins, and 0.50 (IQR 0.50 to 0.51) for distinguishing ACCORD from ACCORD_SPRINT_ Twins, compatible with the SPRINT twins not being distinguishable from the original trial. When assessing differentiation capability for each covariate by training and testing single variate logistic regression models, the overall median accuracy was 0.50-0.51 across comparisons (Figure S3).

### Sensitivity Analyses

In a sensitivity analysis generating the SPRINT_ACCORD_-Twin, we assessed the convergence of the models at various training sample sizes, batch sizes, and number of epochs of training. At a training size of 1% and 25% of the SPRINT data, 81% of the models converged, while 94% converged with 50% and 100% of the data. All models were balanced across the training sample sizes. The proportion of models that reproduced the non-significant hazard ratio seen in ACCORD increased with the training sample and was in a majority of the samples with a sample size >10% of the trial (Table S8).

### Comparison of RCT-Twin-GAN to other Synthesizer Models

Our method consistently scored among the best in all statistical comparisons and correlations (Table S9, S10). It was superior to the other methods in machine learning efficacy, in which a gradient boosting classifier trained on generated digital twin values predicted original RCT values (Table S11).

### Estimating the Primary Composite Outcome in the Non-Conditioned Twins

We confirmed the differences in the reported primary composite outcomes in the SPRINT and ACCORD trials, which included a significant reduction in cardiovascular events in SPRINT’s intervention arm compared with control (hazard ratio 0.75 [0.64-0.89 95% CI, p<0.001]) without a significant reduction in a similar primary composite outcome in ACCORD (hazard ratio 0.88 [0.73 to 1.06 95% CI, p=0.20]). In the SPRINT-Twin without conditioning, the median hazard ratio across 10 generated SPRINT-Twin datasets was 0.73 (CI 0.61-0.87), with the 10 replications performed to ensure the reproducibility of the findings. This was comparable to the HR of 0.75 in the SPRINT trial.^13^ Similarly, the ACCORD-Twin without conditioning replicated the primary results of the ACCORD trial, with a median HR of 0.89 (CI 0.79-1.0) comparable to the HR of 0.88 of the ACCORD trial.^14^

### Estimating the Primary Composite Outcome in the Conditioned Twins

We then demonstrated the ability of RCT-Twins to replicate the known treatment effects of a second population with the SPRINT_ACCORD_-Twin – the SPRINT-Twin that was conditioned on ACCORD. We found the median hazard ratio of 10 SPRINT_ACCORD_-Twin datasets was 0.87 (CI 0.68-1.13), this time comparable to the HR of 0.88 of the ACCORD trial (Figure 4a). In contrast, in 10 replicated digital twins of the ACCORD cohort conditioned on covariate distributions in SPRINT (ACCORD_SPRINT_-Twin), reproduced the significant effect size seen in SPRINT (HR 0.75) with a median hazard ratio of 0.79 (CI 0.72-0.86) (Figure 4b).

**Figure 4:**
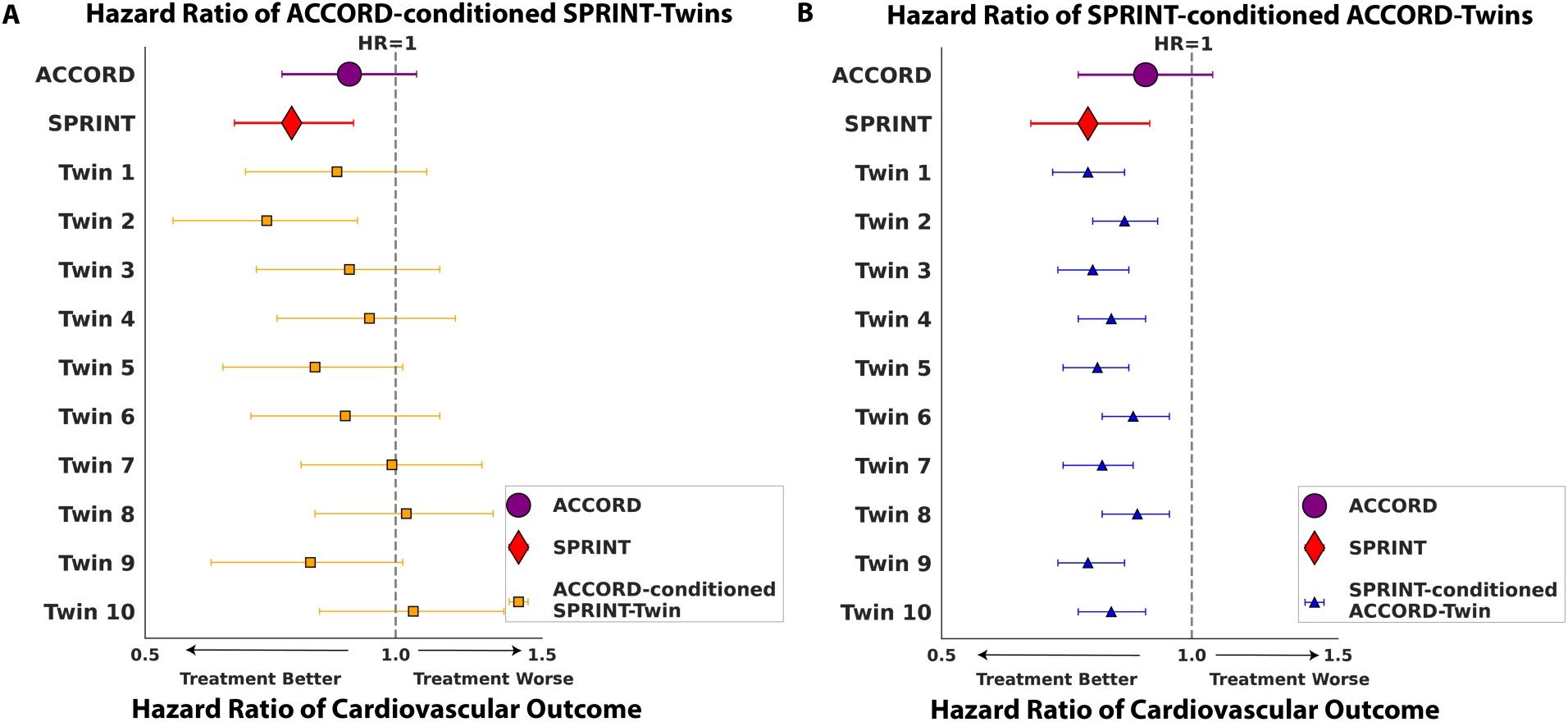
Hazard ratios of intensive blood pressure lowering on cardiovascular outcomes. (A) Forest plot of the SPRINT_ACCORD_-Twin datasets and (B) Forest plot of the ACCORD_SPRINT_-Twin datasets. In both graphs, the purple circle is the ACCORD hazard ratio and 95% confidence interval, the red diamond is the SPRINT hazard ratio and 95% confidence interval, and the grey dotted line represents a hazard ratio of 1. In (A), orange squares are the hazard ratio of major cardiovascular outcome predicted for each twin run of ACCORD-conditioned SPRINT twins with 95% confidence intervals and in (B) the blue triangles are SPRINT-conditioned ACCORD twins. Abbreviations: MACE: Major Cardiovascular Outcomes, SPRINT_ACCORD_-Twin: ACCORD-conditioned SPRINT Twin, ACCORD_SPRINT_-Twin: SPRINT-conditioned ACCORD twin.

**Figure 5:**
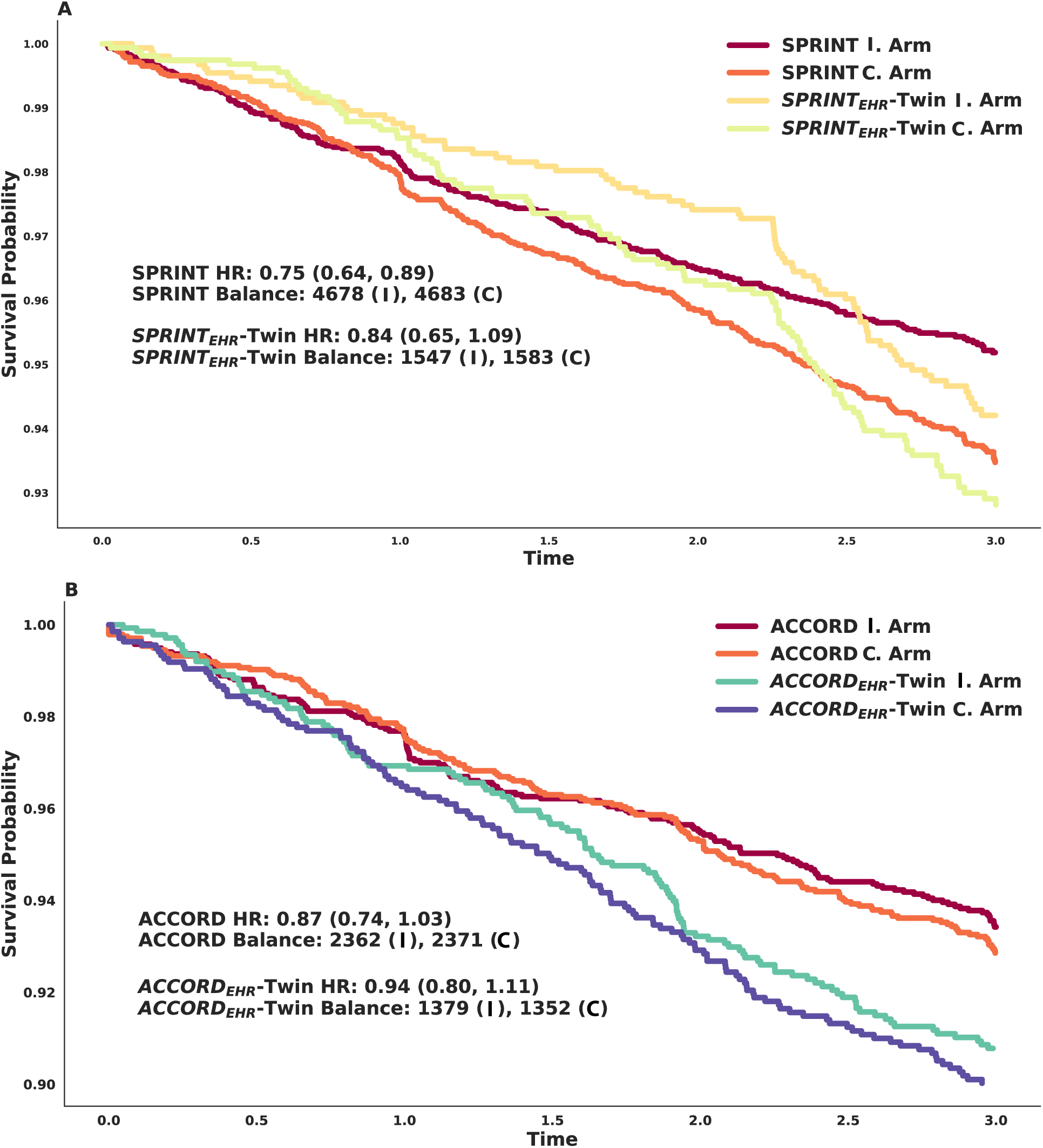
Representative Kaplan-Meier curves of digital twins conditioned on EHR data. (A) Kaplan-Meier curves of SPRINT treatment arms along with EHR-conditioned SPRINT treatment arms, (B) Kaplan Meier curves of ACCORD treatment arms along with EHR-conditioned ACCORD treatment arm balance of the original cohorts and digital twins. Abbreviations: ACCORD_EHR_Twin: ACCORD conditioned on EHR digital twin., C. Control arm, EHR: Electronic Health Record, I: Intervention Arm, SPRINT_EHR_-Twin: SPRINT conditioned on EHR digital twin.

### Estimating the Treatment Effect of SPRINT and ACCORD in the EHR

In a descriptive substudy, we demonstrated the ability to estimate SPRINT and ACCORD primary composite outcomes in patient populations reflecting a large US health system, YNHHS. The same 10 covariates used to build conditioned SPRINT and ACCORD twins were computably extracted from the YNHHS EHR by clinician experts to define covariates in the corresponding EHR cohorts and build digital twins of SPRINT and ACCORD conditioned on corresponding EHR cohorts (Tables S12-S15). In the digital twin of SPRINT conditioned on the corresponding EHR cohort (SPRINT_EHR_-Twin), we confirmed the replication of RCT features, including covariate balance across treatment arms (MASMD 0.03 (SD 0.03), (Figure S4). In this SPRINT_EHR_-Twin the median primary composite outcome HR was 0.84 (95% CI, 0.64-1.09) across the 10 replications. Similarly, the ACCORD_EHR_-Twin replicated both RCT features and EHR covariate distributions, with a median primary composite outcome HR of 0.94 (CI 0.8-1.1).

## DISCUSSION

We present RCT-Twin-GAN, a deep generative model that utilizes clinical knowledge of covariate relationships to synthesize a digital twin of an RCT with selected covariate distributions from a second population, such as another RCT cohort or a general patient population reflected in an EHR. RCT-Twin-GAN created digital twins that replicate the fundamental feature of RCTs, i.e., balanced covariates across treatment arms, but also reflected the covariate distributions of this second population’s distribution. In addition, the RCT-Twin-GAN digital twin cohorts were indistinguishable from the SPRINT RCT cohorts, reproduced RCT covariate correlations, and outperformed other model architectures. Moreover, in a positive control experiment within a two-RCT system where treatment effects were known from well-conducted experiments but were discordant across the RCTs, the RCT-Twins conditioned on covariates from the opposing RCT replicated the results observed in the other RCT, demonstrating the value of examining the effect of population characteristics on study outcomes. We also demonstrate that the approach is flexible to these characteristics drawn from any population, thereby enabling a quantitative evaluation of an RCT’s potential treatment effects in populations that differed from those included in the trial.

Our work has built upon the established need to quantify generalizability of RCTs to new populations.^32^ Prior methods, such as standardization of event rates, allow adjustment by single variables, which groups patients together by singular stratification.^33^ Others have used distance metrics and decision tree machine learning techniques to represent the complex interplay of covariates and characterize the heterogeneity of treatment effect.^8–10,23,34–36^ Prior generative methods have used statistical machine learning to build digital twins of control patients in neurological clinical trials and observational studies with accurate reproduction of patient trajectory at the individual level.^35–37^ Our method complements these by building trial-level digital twins of a conditioning cohort that draw from the multiple covariate distributions and outcomes of an RCT population to generate equivalent covariates in the conditioning population and estimate population-level treatment effects from the RCT intervention. CTAB-GAN+ has been used to build an RCT control patient population, but our studies have demonstrated superiority with DATGAN in reproducing trial baseline characteristics.^38^ In addition, compared to other GAN conditioning methods, our architecture is the only method that can condition on multiple continuous and categorical variables, allowing for multi-variate correlations of the conditioning cohort to be preserved. Statistical methods to assess heterogeneous treatment effects across populations have generally focused on equalizing baseline characteristics between populations using propensity score matching, but this scores one variable at a time, thereby ignoring multi-variable differences across patients, and does not consider effect modifiers.^39^

We incorporate the distributions of multiple mutual pre-randomization covariates available across datasets to ensure representation across multivariate axes. In addition, we utilize clinician expertise to identify connections between covariates and build digital twins modeling the complex interplay of effect modifiers and outcomes. The result is a data-driven generated outcome of the conditioning cohort based on the correlations between multiple covariates and within overlapping covariate distributions between the two patient populations. In Table 1, we discuss the minimum requirements to estimate treatment effects across two populations, including cohort requirements, randomization, intervention and outcome, and sample size.

**Table 1:** Minimal Requirements for RCT-Twin-GAN.

| Rules |
| --- |
| 1. The model requires at least a pair of cohorts, one should be an RCT, the second should have the same covariates accessible and overlapping covariate distributions. This is, however, extendable to any number of cohorts.<br>2. There must be at least two treatment arms in the RCT with any ratio of randomization.<br>3. A measured outcome, categorical or continuous, should be available in the RCT to estimate treatment effect.<br>4. A sample size of at least 100 is needed for model convergence, but at least 1000 participants and further hyperparameter tuning of the model is needed to accurately estimate treatment effects. |

A unique feature of our model incorporates both rigorous statistical methods and clinical knowledge to build digital twins of RCTs with representative covariate balance and effect modifier information. The DAG structure weights clinically relevant relationships between covariates and outcomes and removes spurious correlations that would otherwise be included in the GAN. Of note, the choice of the covariates was governed by primary analysis focused on shared covariates between SPRINT and ACCORD. In real-world translations, a different covariate set shared between a development and target population can be selected. In addition, the ability of the ciDATGAN architecture to condition on multiple continuous and categorical values is unique compared to competing architectures. Our ability to reproduce treatment effect estimates from the conditioning cohort by sampling its covariate distributions relies on the inference of important correlations between covariates during GAN training and digital twin generation. Although prior digital twin studies have focused on supplementing RCTs with synthetic patients for controls^35–38^ and reproducing progression within the same cohort, our study builds upon these by estimating the treatment effect across different patient populations. Measuring the hazard ratios of treatment effect outcomes as an evaluation metric provided valuable insights into the fidelity of the synthetic dataset in simulating clinical trial outcomes and treatment responses.

Methodologically, ACCORD represents a second RCT that experimentally tested the same intervention as SPRINT but in a different population. This is essential as the effect estimates in a conditioned twin otherwise have no gold standard comparison. We demonstrate that conditioning generates effect estimates replicated in the trial that experimentally tested the intervention but arrived at a different conclusion, suggesting the validity of the produced estimates. This is the key methodological outcome of our experiments.

Moreover, there are direct clinical implications for both hypertension management and evidence generation via clinical trials. Our work demonstrates that the observed effect estimate differences in SPRINT and ACCORD emerge because of the nature of populations enrolled in these trials, and not because of diabetes status. There has been a lack of clarity about whether these effects suggested some consideration about blood pressure and its effects on diabetes. But we demonstrate that even in a trial like SPRINT, if the enrolled population had key features that resemble those seen in ACCORD, the trial could have produced a potentially null result. Similarly, had ACCORD enrolled patients that resembled SPRINT – based on features other than diabetes, it could have been positive. While these observations represent data experiments, this was recently observed in the ESPRIT trial, where patients with diabetes benefitted from intensive blood pressure lowering.^40^

This has implications for the interpretation of clinical trials as well. We acknowledge that our work is a proof-of-concept, but we demonstrate that trials can be evaluated in populations that differ from those enrolled on key features to address whether embedded heterogeneous treatment effects and differences in these covariates affect how these results should be interpreted. Moreover, these experiments can guide the need for populations ideally chosen for additional trials. Health systems could determine the likely treatment effect of an intervention in their patient population to better contextualize their patient outcomes with the intervention by developing population-wide digital twins. This effort to use general real-world evidence to establish the efficacy of interventions has major regulatory support from agencies such as the US Food and Drug Administration.^41^

There are limitations to consider. First, RCT-Twin-GAN uses a select set of variables to build the digital twin. We chose a smaller set of covariates to maximize efficiency and showed that even with this small number of representative variables, we can build a digital twin that successfully replicates treatment effect estimates. Second, our model relies on outside input for identifying correlations between covariates, but we believe this can be considered a strength that clinical expertise can be imbued into the model to reduce the weight of spurious correlations inherent in data. Third, this is a post-hoc analysis of RCTs, but we show the ability of digital twins to mirror covariate characteristics and treatment effects found in SPRINT and ACCORD. Fourth, we only applied RCT-Twin-GAN to the SPRINT – ACCORD pair because it was the only paired trial testing the same intervention with different results available through a public domain, the National Heart, Lung, and Blood Institute Biologic Specimen and Data Repository Information Coordinating Center (BioLINCC). As the data are publicly available, further research can build upon this example, and we further anticipate applying our model to other examples.

Fifth, we could not study glycemic effects on the intervention because SPRINT did not control hyperglycemia or include diabetic patients as seen in ACCORD. Despite this, we demonstrate a positive treatment effect aligned with SPRINT among the ACCORD patients, who had a full range of glycemic management differences as part of the original ACCORD trial, and so do not find evidence to suggest glycemic management differences produced the null results observed in ACCORD. Sixth, GANs are known to have challenges with achieving successful convergence between the discriminator and generator, so we have adapted the most successful advances in GAN development to optimize convergence. We chose an architecture that reduces spurious correlations by introducing a DAG to define the correlation structure between variables, stabilized training with the most appropriate learning rate and batch normalization layers, and incorporated a loss function that eliminated the risk of vanishing gradients to ensure optimal model performance. Seventh, modeling real-world patients in the EHR can be challenging since the data represents a snapshot of patients who seek care, but we choose patients from a diverse tertiary care system to maximize the breadth of the general population identified. In addition, the EHR covariates had to be operationally defined by experts to be analogous to the criteria used in RCT, but this is a descriptive study that shows different covariate distributions can be modeled. Finally, the true effect estimates in the EHR populations are unknown, and those estimated by RCT-Twin-GAN should not inform care but rather give an idea of discordance or concordance with the original RCT population.

We have introduced a new application of GANs to build synthetic cohorts by creating an RCT digital twin reflective of different patient populations, including similar RCTs and real-world patients found in the EHR. Our study demonstrates a way to evaluate the generalizability of an RCT to the general population by embedding covariate distributions that are more representative of real-world populations. This amplifies the effects for those more frequently seen in clinical practice. Overall, our model contributes significantly to the evidence supporting the development of an RCT digital twin that more consistently mirrors real-world populations, thereby enhancing inference for real-world patients.

## METHODS

### Data Source and Patient Populations

#### SPRINT and ACCORD Cohorts

From 2010-2013, at 102 clinical sites across the United States, participants were recruited for the SPRINT RCT who were at least 50 years old, had a systolic blood pressure between 130 and 180 mm Hg, and had increased cardiovascular event risk, including cardiovascular disease with the exception of stroke, chronic kidney disease, Framingham 10 year cardiovascular risk score of 15% or greater, and advanced age over 75. Patients with prior stroke, diabetes mellitus, and a recent heart failure exacerbation had been excluded from the study.

From 2001 to 2005, at 77 clinical sites across the United States and Canada, participants were recruited for the ACCORD RCT who had type 2 diabetes mellitus, a glycated hemoglobin level of 7.5% or greater, and either age 40 or older with cardiovascular disease or age 55 or older with risk factors for cardiovascular disease and anatomical evidence of longstanding hypertension or diabetes such as albuminuria or left ventricular hypertrophy. Patients with a BMI over 45, a creatinine over 1.5 mg/dL, or serious illness were excluded.

#### EHR cohorts

The two EHR cohorts were extracted from patients within the Yale New Haven Health System (YNHHS) from 2013 to 2023. The study was reviewed by the Yale Institutional Review Board and deemed exempt as it uses retrospective data. We sampled 100,000 adult patients and then filtered the cohort to those with an ICD-10-CDM code for hypertension (Table S16). Out of these patients, we filtered for patients with an ICD-10-CDM code for type 2 diabetes mellitus (Table S16). Patients with both hypertension and type 2 diabetes mellitus billing codes were considered for the ACCORD EHR cohort. The remaining hypertension patients who did not have type 2 diabetes mellitus billing codes were considered for the SPRINT EHR cohort. We excluded patients who did not have values for continuous covariates and patients above the age of 110. We then sampled 4000 patients each for the ACCORD EHR and SPRINT EHR cohorts with values for all conditioned covariates. We further excluded patients who had continuous values out of range of the training cohort of SPRINT or ACCORD (Table S17).

### Development of RCT Digital Twins Conditioned on a Second Patient Population

We adapted CGAN models to create digital twin datasets of an RCT conditioned on covariate distributions from a second patient population. We first built a SPRINT digital twin (SPRINT-twin) trained on the SPRINT cohort without a second conditioning cohort. We then built a SPRINT digital twin conditioned on the ACCORD participant population (SPRINT_ACCORD_-Twin) with the intention of reproducing the ACCORD primary composite outcome in a SPRINT digital twin (Figure 1). To implement this, we applied the Conditional inputs for Direct Acyclic Tabular Generative Adversarial Networks (CiDATGANs), a conditional tabular GAN that uses a directed acyclic graph (DAG) to assign relationships between pre-randomized covariates.^29,42^ The DAG ensures clinically relevant connections are introduced between covariates and prevents the weighting of spurious correlations between covariates. To condition the digital twins on the other RCT population, we mapped 33 equivalent covariates between SPRINT and ACCORD (Table S18).

### Covariate Extraction for SPRINT, ACCORD, and the EHR

In order to condition the SPRINT digital twin (SPRINT-Twin) on equivalent ACCORD covariates (SPRINT_ACCORD_-Twin), we mapped 33 equivalent covariates between the two cohorts, which included demographics such as age, gender, race, and ethnicity, conditions and social history, such as smoking history, family history of cardiovascular disease (CVD), hyperlipidemia, left ventricular hypertrophy (LVH) and prior myocardial infarction (MI), medications such as taking aspirin or statins, procedures such as coronary revascularization, and laboratory values and vital signs such as glomerular filtration rate (GFR), glucose, and systolic blood pressure (Table S18). We also included outcome, time to outcome, and treatment arm assignment. We limited the maximum time to outcome to five years, censoring all subsequent outcomes.

To build the DAG, an expert clinician identified 16 representative variables of the 33 mapped between SPRINT and ACCORD to represent all clinical areas such as demographics, conditions, medications, family history, symptoms, social history, procedures, vital signs, and laboratory and EKG measures, and also maintaining a balance of both categorical and continuous variables. All demographic variables were included since they are available for everyone in the EHR cohort. The variables included continuous covariates of age at randomization, GFR, heart rate, LDL cholesterol, and systolic blood pressure, and categorical covariates converted to a binary assignment of the presence (1) or absence (0) of angina, Black race, BMI, current smoker, family history of CVD, female sex, Hispanic ethnicity, LVH, previous MI, statin use, and White race (Table S18). Since BMI was considered a binary variable in ACCORD (above or below 32 kg/m^2^), we used a similar definition in SPRINT.

Variables related to exclusion criteria of at least one of the cohorts were not included in the conditioning of the model or constructing the DAG because of the lack of overlap in the distribution of these covariate values between the SPRINT and ACCORD cohorts. These included glucose and diabetes mellitus. The DAG construction includes an iterative process of expert assessment of clinically relevant pairs and the causal direction within the pairs and calculation of correlations between unpaired variables (Table 2). The final DAG included 71 connections (Figure 2, Table S19). The arrows’ direction pointed from the independent covariate to the dependent covariate. No arrow pointed to the treatment arm covariate, labeled “Group”, since this assignment was independent of all covariates. All covariates and the “Group” pointed to the “Outcome” and “Time to Outcome” covariates since all covariates and treatment arm assignment were thought to influence the outcome (Figure 2, Table S19). We used the 10 covariates with the largest absolute standardized mean difference between SPRINT and ACCORD as the conditioned covariates in order to condition from the covariate distributions most representative of the second cohort. The included binary and continuous covariates, in the order of increasing dissimilarity between cohorts, were black race, history of previous MI, female sex, statin use, LVH, BMI, heart rate, age at randomization, family history of CVD, and GFR.

**Table 2:**
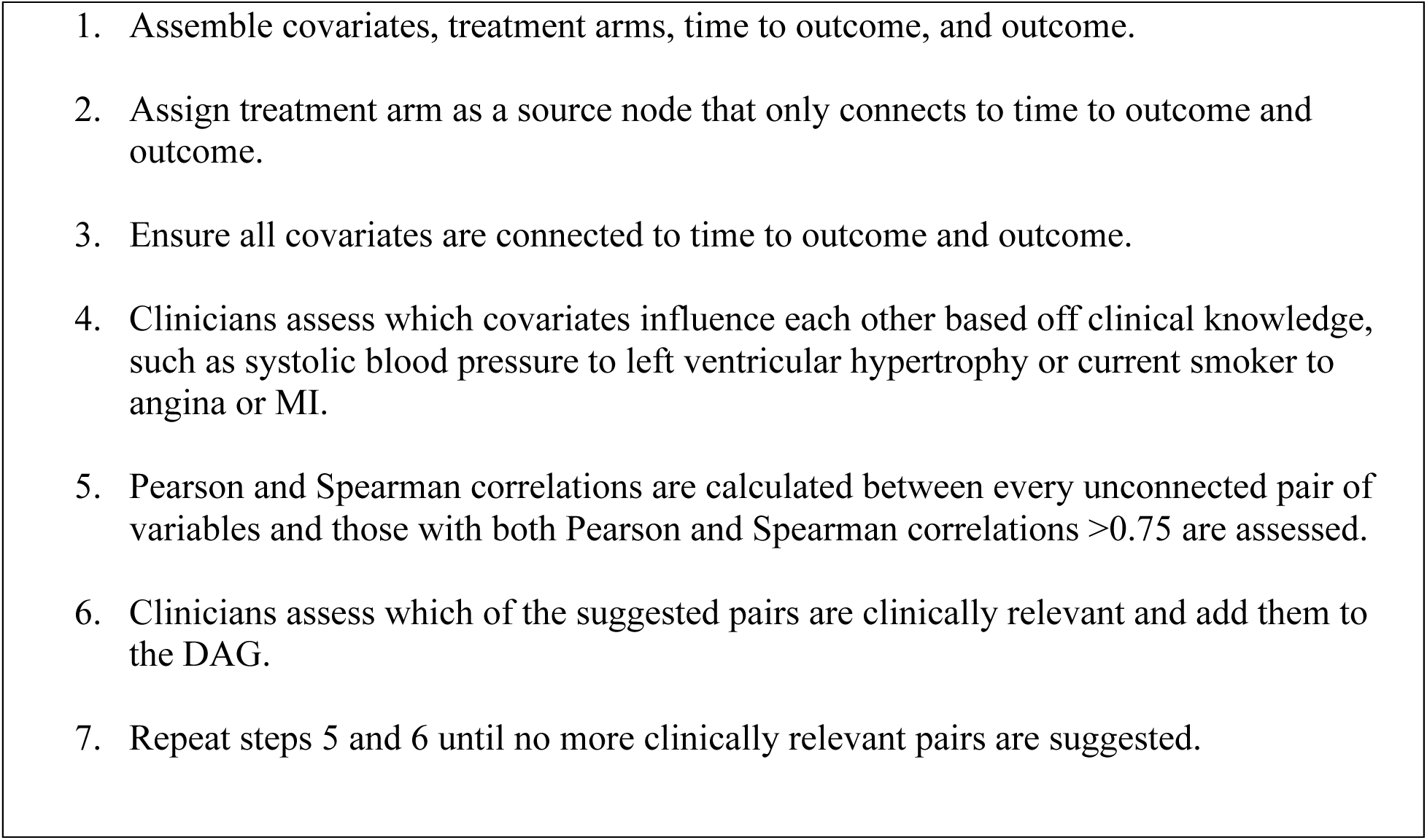
Construction of the Directed Acyclic Graph.

**Table 3:**
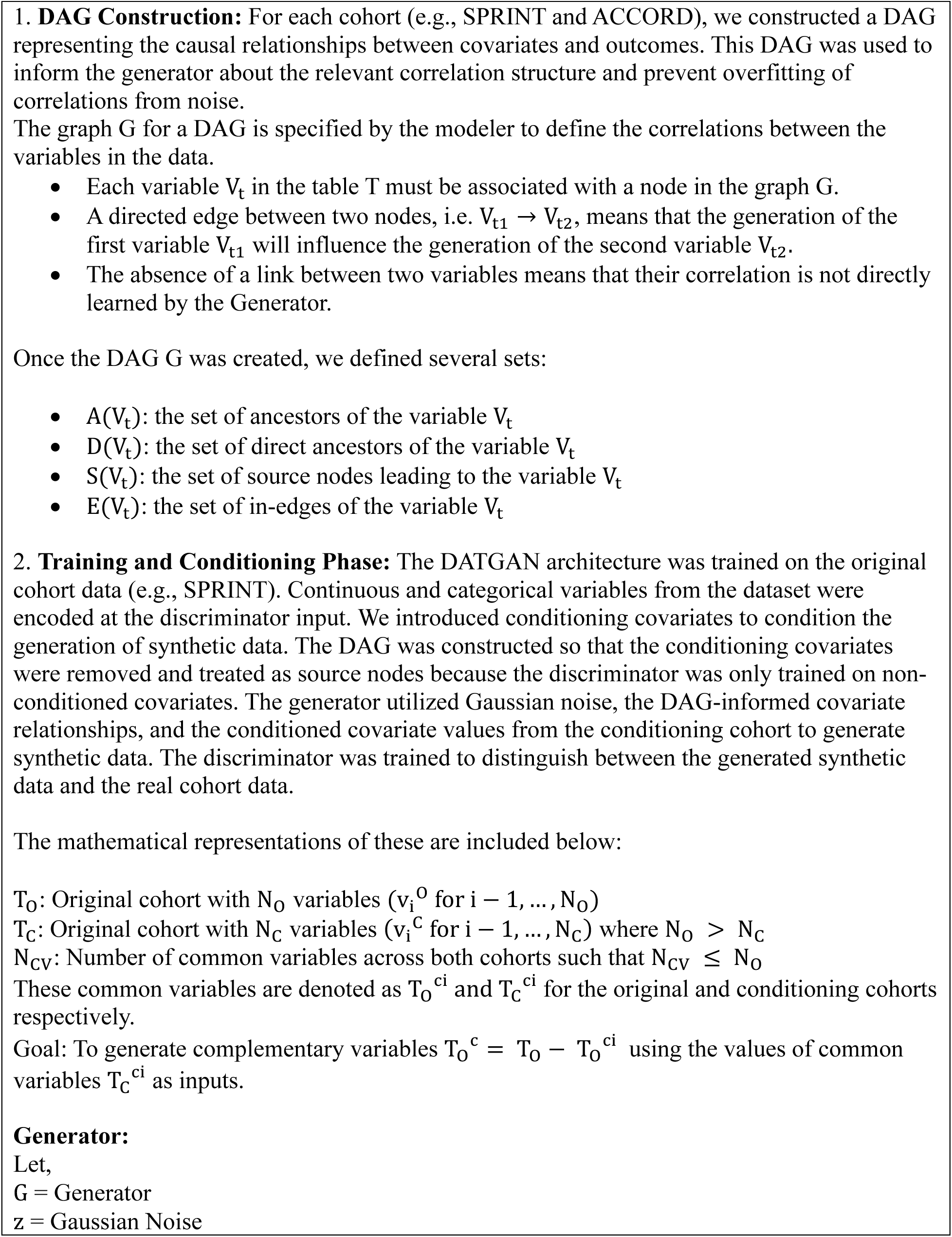
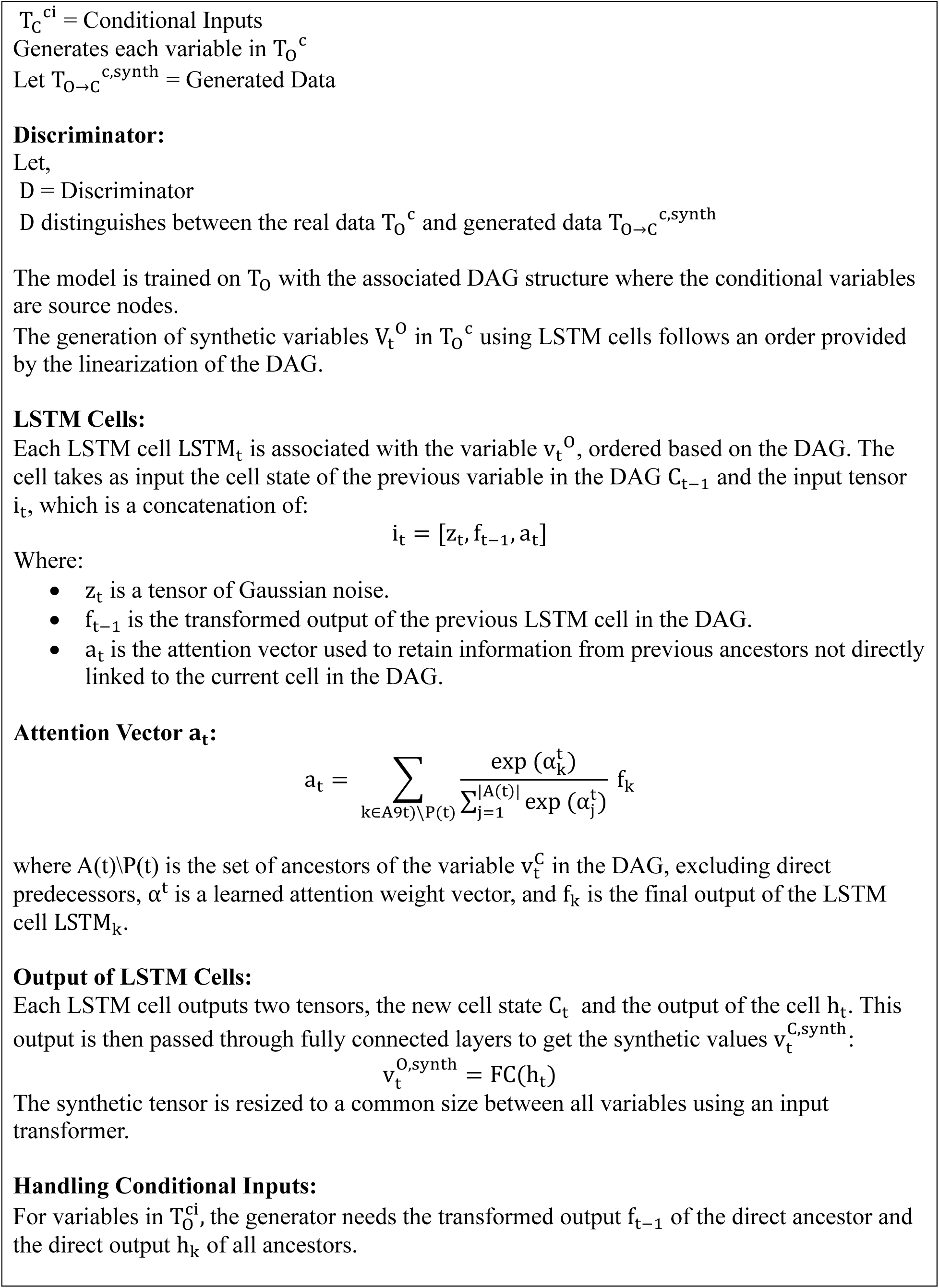
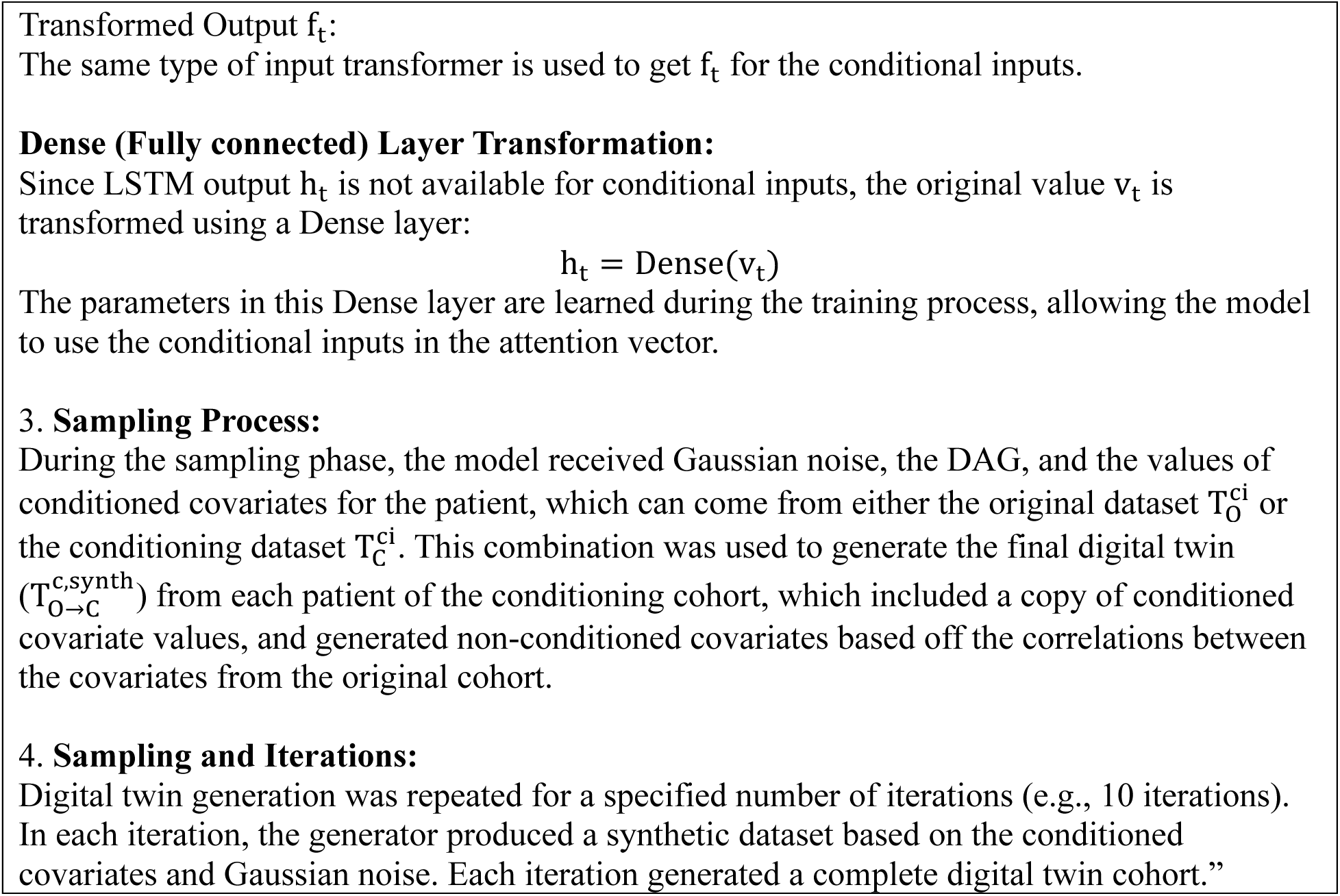
CiDATGAN Architecture, Training and Sampling.

Since we sought to condition on the EHR populations as well, we extracted the 10 conditioned covariates established in the prior analysis from the EHR as well (Table S16). Only patients with a value for the demographics sex, race, and age (based on an available date of birth), the vital signs BMI and heart rate, and the laboratory test eGFR (or computable from serum creatinine), were included. The binary covariates of family history of CVD, LVH, previous MI, and statin use were considered not present (assigned 0) if they were not recorded in the patient’s EHR, as is the norm for observational research studies in the EHR.^43^ Age was calculated on October 1, 2023 (EHR query date), unless they were deceased, where we used the death date to define their last known age. We used this index date to consider most current clinical characteristics of the patient to estimate their treatment effect, the equivalent of the randomization to treatment arm date in the RCT.

### Design of the RCT-Twin-GAN Model

RCT-Twin-GAN is a Generative Adversarial Network model, which is a deep learning model rooted in game theory that pits a generator, the neural network that creates synthetic data, against a discriminator, the neural network that distinguishes between the real data it is trained on and the synthetic data created by the generator. The minimization of the discrimination between real and synthetic data allows for the GAN to make realistic digital twins of the cohort on which it is developed.^44^ The neural networks are comprised of Long Short Term Memory (LSTM) cells, which are structured to retain information from prior inputs in addition to the current variable input.^45^ GANs have been adapted to accurately synthesize tabular data such as EHR data.^27,28,46^ GANs can also integrate data from a second patient population through conditioning the model on sample covariate values from the original cohort within the covariate distribution of the conditioning cohort, or the second patient population.^28,29,46^ To avoid the well-documented challenges with consistently achieving convergence in GANs, our model utilizes Wasserstein loss to overcome training instability and prevent vanishing gradients.^47^

RCT-Twin-GAN is based on the architecture of CiDATGAN, which is an extension of DATGAN with an additional feature of conditioning covariates with distributions from a second population.^29,42^ The DATGAN and CiDATGAN models employ a unique feature, allowing the generator to have the relationships between covariates and outcomes of the original training cohort to be explicitly encoded via a Directed Acyclic Graph (DAG). This prevents overfitting of the discriminator by defining the correlation structure between variables. It constrains the number of relevant associations, prioritizing key features for the model to learn. In contrast to other conditional GAN architectures that infer variable relationships solely by correlation, the addition of DAG to the training of the CiDATGAN generator incorporates directed relationships between pairs of variables to eliminate spurious correlations between variables. Continuous variables were winsorized based on the min-max values of the covariate in the training dataset (Table S17) to remove outlier values below the 2.5% and 97.5% percentiles, and categorical variables were encoded into one-hot vectors and then fed into the discriminator as part of the input.

During the training phase, the generator combines Gaussian noise and attention vectors of the LSTM cells in the order of the DAG relationships and transforms the covariates from the original cohort using a fully connected layer in order to refine relationships and dependencies between the inputs. CiDATGAN creates a key modification to the DATGAN architecture in which the transformed conditional covariate inputs are also fed to the generator. Because of this modification, the DAG is also modified so that all conditional covariates are source nodes. The generator then synthesizes complementary values of the remaining covariates of the original dataset. The discriminator is then trained to differentiate between the original versus generated values of the remaining covariates from the original dataset. The discriminator is then trained to differentiate between the original versus generated values of the remaining covariates from the original dataset (Figure 1a).

During the sampling phase, the generator receives Gaussian noise, the modified DAG, and conditioned covariate values from the conditioning cohort and produces synthetic data without transformation in order to directly reflect the learned distribution from training while maintaining the integrity of the inputs from the conditioning cohort. Therefore, the final synthetic dataset incorporates the conditioning cohort inputs and generates complementary values for the remaining covariates missing from the conditioning cohort. The generator creates a cohort digital twin by producing one row of data at a time for each patient within the conditioning cohort (Figure 1b).

The CiDATGAN was then trained with the DAG and encoded dataset to generate the synthetic dataset. We performed a hyperparameter grid search with different batch sizes and epochs to find the best parameters to generate synthetic data with similar outcomes to the original dataset (Table S20).

### Application of RCT-Twin-GAN in SPRINT, ACCORD, and the EHR

We first used RCT-Twin-GAN to build a SPRINT-Twin, which created a DAG based on the SPRINT cohort, trained the DATGAN architecture on the SPRINT cohort, and ran the sampling phase of the DATGAN pipeline ten times, resulting in ten distinct synthetic twins. We then built a SPRINT_ACCORD_-Twin, which again created a DAG from and trained on the SPRINT cohort but was conditioned on the ACCORD cohort, utilizing the CiDATGAN architecture. This meant that the DAG was modified to remove connections going to the conditioning covariates, and in the sampling phase, Gaussian noise and the conditioned covariate distributions from the ACCORD cohort were inputs for the generator in order to create the final synthetic dataset. We sampled this process for 10 iterations to make 10 SPRINT_ACCORD_-Twin datasets. We repeated this process to create ACCORD_SPRINT_-Twin datasets by replacing ACCORD to be the training cohort and SPRINT to be the conditioning cohort.

We repeated this training and conditioning process using the EHR cohorts as well. Specifically, we trained RCT-Twin-GAN on the SPRINT cohort, conditioned on the SPRINT-EHR cohort, and sampled 10 times to create SPRINT_EHR_-Twins. We similarly made ACCORD_EHR_-Twins with ACCORD and the ACCORD-EHR cohort.

### Analysis of Cohort Representation in Digital Twins

To determine whether RCT-Twin-GAN created digital twins that are balanced by treatment arm, we calculated the mean absolute standardized mean difference (MASMD) of all covariates of the digital twins stratified by treatment arm assignment. A value of less than 0.1 was considered adequate balance, consistent with convention when assessing the success of propensity score matching.^48^ To assess the representation of the conditioning cohort in the synthetic digital twin, we also calculated the absolute standardized mean difference (ASMD) between SPRINT and ACCORD for each covariate, SPRINT-Twin and ACCORD, and SPRINT_ACCORD_-Twin and ACCORD. We also calculated the Spearman correlation between all variables for each RCT and digital twin, discretized the correlations into 7 bins from –1 to 1, and then calculated the proportion of covariate correlations from the RCT and twin data that were in the same bin (termed correlation accuracy) and mean absolute difference between RCT covariate correlation values and digital twin covariate correlation values as described in Li et al.^27^

### Evaluation of Digital Twin Similarity and Integrity

We evaluated whether any row of the digital twin cohorts matched the RCT patients by a similarity score evaluation by Synthetic Data Vault.^49^ To assess distinguishability of the RCT data from the conditioned twins, we trained and tested with a 70/30 split a multivariable logistic regression classifier to differentiate between the RCT and digital twin, which has been used to assess the integrity of prior digital twins.^37^

### Comparison of RCT-Twin-GAN to other synthesizer models

We compared the DATGAN architecture used in RCT-Twin-GAN to 5 competing models including Conditional Tabular GAN (CTGAN)^28^, Conditional Tabular GAN+ (CTABGAN+)^50^, CopulaGAN^51^, GaussianCopula^51^, and Triplet-based Variational Autoencoder (TVAE)^28^. We utilized the non-conditioned DATGAN architecture because none of the comparator models have the flexibility to condition multiple continuous and categorical variables like the CiDATGAN architecture. Specifically, we tested the mean absolute error, R^2^, root mean squared error, standardized root mean squared error, and Pearson correlation between the distribution of unique values in the RCT compared to the digital twin. We also tested machine learning efficacy, in which a gradient bosting classifier is trained on twin data and evaluated on how well it generates real data.

### Estimation of Treatment Effect on Cardiovascular Outcomes in the Digital Twins

In order to assess the ability of RCT-Twin-GAN to estimate RCT treatment effect outcomes in populations other than the original RCT, we calculated the hazard ratio of cardiovascular outcomes stratified by treatment arms in each of the digital twin cohorts using cox proportional hazard models. We utilized hazard ratios to evaluate the comparative risks of events over time between different treatment groups within the synthetic data. This analytical approach allowed us to gauge the effectiveness of the synthetic dataset in accurately representing the underlying dynamics of treatment effects and event occurrences observed in real-world scenarios.

We reported the median hazard ratio and 95% confidence intervals for the 10 SPRINT-Twin, SPRINT_ACCORD_-Twin, and ACCORD_SPRINT_-Twin digital twins. In order to demonstrate the ability to estimate treatment effect outcomes in a variety of cohorts, we calculated the hazard ratio and 95% confidence intervals of cardiovascular outcomes of the SPRINT_EHR_-Twins and ACCORD_EHR_-Twins as well.

### Statistical Analysis

Categorical variables were summarized as numbers with percentages, and continuous variables were summarized as median with 25% and 75% interquartile ranges (IQR) or mean with standard deviation (SD). Covariate distributions were compared using ASMD and standard deviations and Spearman correlations between covariates and graphed as love plots comparing datasets and heatmaps of the correlations. Data was winsorized at 2.5% and 97.5% percentiles to remove outliers. Survival analysis was conducted using unadjusted cox proportional hazard models with p values calculated after 5 years and presented as Kaplan Meier survival curves.

Hazard ratios across digital twins and SPRINT and ACCORD cohorts were presented as forest plots with 95% confidence interval error bars. Analyses were conducted using python 3.9, with packages specified in the supplement.

## DATA AVAILABILITY

The SPRINT and ACCORD cohorts are publicly available through the National Heart, Lung, and Blood Institute Biologic Specimen and Data Repository Information Coordinating Center (BioLINCC). The SPRINT dataset is available at https://biolincc.nhlbi.nih.gov/studies/sprint/ and the ACCORD dataset is available at https://biolincc.nhlbi.nih.gov/studies/accord/. The Yale electronic health record cohorts are not available due to the use of patient data.

## CODE AVAILABILITY

The code for reproducing the treatment effect estimates, digital twins, and analysis figures will be available during peer review in an accompanying file, and the code will be made publicly available upon publication.

## ACKNOWLEDGEMENTS

The study is supported by the National Heart, Lung, and Blood Institute of the National Institutes of Health (R01HL167858). Dr. Thangaraj and Dr. Oikonomou are also supported by the National Heart, Lung, and Blood Institute of the National Institutes of Health (5T32HL155000-03 and 1F32HL170592-01, respectively).

## CONTRIBUTIONS

PMT and SVS contributed equally to the study. RK conceived the study and PMT, SVS, EKO, and RK drafted a research plan. EKO and PMT accessed and processed the data. PMT and SVS developed and analyzed the GAN model. PMT, SVS, EKO, and RK drafted the manuscript. All authors provided feedback regarding the study design and manuscript. RK supervised the study, procured funding, and is the guarantor.

## REFERENCES

1. Averitt AJ, Ryan PB, Weng C, Perotte A. A conceptual framework for external validity. J Biomed Inform. 2021;121:103870.

2. Rothwell PM. External validity of randomised controlled trials: “To whom do the results of this trial apply?” Lancet. 2005;365:82–93.

3. Filbey L, Zhu JW, D’Angelo F, et al. Improving representativeness in trials: a call to action from the Global Cardiovascular Clinical Trialists Forum. Eur Heart J. 2023;44:921–930.

4. Kennedy-Martin T, Curtis S, Faries D, Robinson S, Johnston J. A literature review on the representativeness of randomized controlled trial samples and implications for the external validity of trial results. Trials. 2015;16:495.

5. Ranganathan M, Bhopal R. Exclusion and inclusion of nonwhite ethnic minority groups in 72 North American and European cardiovascular cohort studies. PLoS Med. 2006;3:e44.

6. Sardar MR, Badri M, Prince CT, Seltzer J, Kowey PR. Underrepresentation of women, elderly patients, and racial minorities in the randomized trials used for cardiovascular guidelines. JAMA Intern Med. 2014;174:1868–1870.

7. DeFilippis EM, Echols M, Adamson PB, et al. Improving Enrollment of Underrepresented Racial and Ethnic Populations in Heart Failure Trials: A Call to Action From the Heart Failure Collaboratory. JAMA Cardiol. 2022;7:540–548.

8. Oikonomou EK, Spatz ES, Suchard MA, Khera R. Individualising intensive systolic blood pressure reduction in hypertension using computational trial phenomaps and machine learning: a post-hoc analysis of randomised clinical trials. Lancet Digit Health. 2022;4:e796–e805.

9. Oikonomou EK, Suchard MA, McGuire DK, Khera R. Phenomapping-Derived Tool to Individualize the Effect of Canagliflozin on Cardiovascular Risk in Type 2 Diabetes. Diabetes Care. 2022;45:965–974.

10. Oikonomou EK, Van Dijk D, Parise H, et al. A phenomapping-derived tool to personalize the selection of anatomical vs. functional testing in evaluating chest pain (ASSIST). Eur Heart J. 2021;42:2536–2548.

11. Patel HC, Hayward C, Dungu JN, et al. Assessing the Eligibility Criteria in Phase III Randomized Controlled Trials of Drug Therapy in Heart Failure With Preserved Ejection Fraction: The Critical Play-Off Between a “Pure” Patient Phenotype and the Generalizability of Trial Findings. J Card Fail. 2017;23:517–524.

12. Lim YMF, Molnar M, Vaartjes I, et al. Generalizability of randomized controlled trials in heart failure with reduced ejection fraction. Eur Heart J Qual Care Clin Outcomes. 2022;8:761– 769.

13. SPRINT Research Group, Wright JT Jr, Williamson JD, et al. A Randomized Trial of Intensive versus Standard Blood-Pressure Control. N Engl J Med. 2015;373:2103–2116.

14. ACCORD Study Group, Cushman WC, Evans GW, et al. Effects of intensive blood-pressure control in type 2 diabetes mellitus. N Engl J Med. 2010;362:1575–1585.

15. Carson JL, Brooks MM, Hébert PC, et al. Restrictive or Liberal Transfusion Strategy in Myocardial Infarction and Anemia. N Engl J Med. 2023;389:2446–2456.

16. Ducrocq G, Gonzalez-Juanatey JR, Puymirat E, et al. Effect of a Restrictive vs Liberal Blood Transfusion Strategy on Major Cardiovascular Events Among Patients With Acute Myocardial Infarction and Anemia: The REALITY Randomized Clinical Trial. JAMA. 2021;325:552–560.

17. Joosten LPT, van Doorn S, van de Ven PM, et al. Safety of Switching from a Vitamin K Antagonist to a Non-Vitamin K Antagonist Oral Anticoagulant in Frail Older Patients with Atrial Fibrillation: Results of the FRAIL-AF Randomized Controlled Trial. Circulation. 2023. Published onlineAugust 27, 2023. 10.1161/CIRCULATIONAHA.123.066485.

18. Granger CB, Alexander JH, McMurray JJV, et al. Apixaban versus Warfarin in Patients with Atrial Fibrillation. N Engl J Med. 2011;365:981–992.

19. Jane-wit D, Horwitz RI, Concato J. Variation in results from randomized, controlled trials: stochastic or systematic? J Clin Epidemiol. 2010;63:56–63.

20. Krakoff LR. A tale of 3 trials: ACCORD, SPRINT, and SPS3. What happened? Am J Hypertens. 2016;29:1020–1023.

21. Chobanian AV. Hypertension in 2017-what is the right target? JAMA. 2017;317:579–580.

22. Huang C, Dhruva SS, Coppi AC, et al. Systolic blood pressure response in SPRINT (Systolic Blood Pressure Intervention Trial) and ACCORD (Action to Control Cardiovascular Risk in Diabetes): A possible explanation for discordant trial results. J Am Heart Assoc. 2017;6.

23. Laffin LJ, Besser SA, Alenghat FJ. A data-zone scoring system to assess the generalizability of clinical trial results to individual patients. Eur J Prev Cardiol. 2019;26:569–575.

24. Liu R, Rizzo S, Whipple S, et al. Evaluating eligibility criteria of oncology trials using real-world data and AI. Nature. 2021;592:629–633.

25. Ge Q, Huang X, Fang S, et al. Conditional Generative Adversarial Networks for Individualized Treatment Effect Estimation and Treatment Selection. Front Genet. 2020;11:585804.

26. Yoon J, Jordon J, Van Der Schaar M. Ganite: Estimation of individualized treat-ment effects using generative adversarial nets. 2018. Accessed November 9, 2023. https://openreview.net/pdf?id=ByKWUeWA-.

27. Li J, Cairns BJ, Li J, Zhu T. Generating synthetic mixed-type longitudinal electronic health records for artificial intelligent applications. NPJ Digit Med. 2023;6:98.

28. Xu L, Skoularidou M, Cuesta-Infante A, Veeramachaneni K. Modeling Tabular data using Conditional GAN. arXiv [csLG]. 2019.

29. Lederrey G, Hillel T, Bierlaire M. ciDATGAN: Conditional Inputs for Tabular GANs. arXiv [csLG]. 2022.

30. He Z, Tang X, Yang X, et al. Clinical Trial Generalizability Assessment in the Big Data Era: A Review. Clin Transl Sci. 2020;13:675–684.

31. Tripepi G, Jager KJ, Dekker FW, Zoccali C. Stratification for confounding--part 2: direct and indirect standardization. Nephron Clin Pract. 2010;116:c322–5.

32. Duan T, Rajpurkar P, Laird D, Ng AY, Basu S. Clinical Value of Predicting Individual Treatment Effects for Intensive Blood Pressure Therapy. Circ Cardiovasc Qual Outcomes. 2019;12:e005010.

33. Brantner CL, Nguyen TQ, Tang T, Zhao C, Hong H, Stuart EA. Comparison of methods that combine multiple randomized trials to estimate heterogeneous treatment effects. Stat Med. 2024. Published onlineJanuary 25, 2024. 10.1002/sim.9955.

34. Raghavan S, Josey K, Bahn G, et al. Generalizability of heterogeneous treatment effects based on causal forests applied to two randomized clinical trials of intensive glycemic control. Ann Epidemiol. 2022;65:101–108.

35. Fisher CK, Smith AM, Walsh JR, Coalition Against Major Diseases, Abbott, Alliance for Aging Research, Alzheimer’s Association, Alzheimer’s Foundation of America, AstraZeneca Pharmaceuticals LP, Bristol-Myers Squibb Company, Critical Path Institute, CHDI Foundation, Inc., Eli Lilly and Company, F. Hoffmann-La Roche Ltd, Forest Research Institute, Genentech, Inc., GlaxoSmithKline, Johnson & Johnson, National Health Council, Novartis Pharmaceuticals Corporation, Parkinson’s Action Network, Parkinson’s Disease Foundation, Pfizer, Inc., sanofi-aventis. Collaborating Organizations: Clinical Data Interchange Standards Consortium (CDISC), Ephibian, Metrum Institute. Machine learning for comprehensive forecasting of Alzheimer’s Disease progression. Sci Rep. 2019;9:13622.

36. Walsh JR, Smith AM, Pouliot Y, Li-Bland D, Loukianov A, Fisher CK. Generating Digital Twins with Multiple Sclerosis Using Probabilistic Neural Networks. arXiv [statML]. 2020.

37. Bertolini D, Loukianov AD, Smith AM, et al. Modeling Disease Progression in Mild Cognitive Impairment and Alzheimer’s Disease with Digital Twins. arXiv [csLG]. 2020.

38. Eckardt J-N, Hahn W, Röllig C, et al. Mimicking clinical trials with synthetic acute myeloid leukemia patients using generative artificial intelligence. NPJ Digit Med. 2024;7:76.

39. Degtiar I, Rose S. A Review of Generalizability and Transportability. Annual Review of Statistics and Its Application. 2023;10:501–524.

40. Liu J, Li Y, Ge J, et al. Lowering systolic blood pressure to less than 120 mm Hg versus less than 140 mm Hg in patients with high cardiovascular risk with and without diabetes or previous stroke: an open-label, blinded-outcome, randomised trial. Lancet. 2024;404:245–255.

41. Center for Drug Evaluation and Research (CDER), Center for Biologics Evaluation and Research (CBER) U.S. Food and Drug Administration. Framework for FDA’s Real World Evidence Program. US Food & Drug Administration. 2018. Accessed March 6, 2024. https://www.fda.gov/science-research/science-and-research-special-topics/real-world-evidence.

42. Lederrey G, Hillel T, Bierlaire M. DATGAN: Integrating expert knowledge into deep learning for synthetic tabular data. arXiv [csLG]. 2022.

43. Khera R, Schuemie MJ, Lu Y, et al. Large-scale evidence generation and evaluation across a network of databases for type 2 diabetes mellitus (LEGEND-T2DM): a protocol for a series of multinational, real-world comparative cardiovascular effectiveness and safety studies. BMJ Open. 2022;12:e057977.

44. Goodfellow IJ, Pouget-Abadie J, Mirza M, et al. Generative Adversarial Networks. arXiv [statML]. 2014.

45. Hochreiter S, Schmidhuber J. Long short-term memory. Neural Comput. 1997;9:1735–1780.

46. Zhao Z, Kunar A, Van der Scheer H, Birke R, Chen LY. CTAB-GAN: Effective Table Data Synthesizing. arXiv [csLG]. 2021.

47. Arjovsky M, Chintala S, Bottou L. Wasserstein GAN. arXiv [statML]. 2017.

48. Normand ST, Landrum MB, Guadagnoli E, et al. Validating recommendations for coronary angiography following acute myocardial infarction in the elderly: a matched analysis using propensity scores. J Clin Epidemiol. 2001;54:387–398.

49. Patki N, Wedge R, Veeramachaneni K. The synthetic data vault. In: 2016 IEEE International Conference on Data Science and Advanced Analytics (DSAA). IEEE, 2016.

50. Zhao Z, Kunar A, Birke R, Chen LY. CTAB-GAN+: Enhancing Tabular Data Synthesis. arXiv [csLG]. 2022.

51. Kamthe S, Assefa S, Deisenroth M. Copula flows for synthetic data generation. arXiv [statML]. 2021.

