## Supplementary Information for "A Novel Digital Twin Strategy to Examine the Implications of Randomized Clinical Trials for Real-World Populations"

### Table of Contents

| Title | Page |
|---|---|
| Figure S1: Spearman Correlations between variables within RCTs and Twins | 3 |
| Figure S2: Absolute standardized mean difference (ASMD) between RCTs and digital twins conditioned with the other RCT | 4 |
| Figure S3: Logistic Regression Accuracy of RCT vs Non-conditioned Twin | 5 |
| Figure S4: ASMD between treatment and placebo arms of RCTs and EHR digital twins | 6 |
| Figure S5: Representative Kaplan Meier curves of digital twins conditioned on RCT data | 7 |
| Table S1: Baseline Characteristics of 5 SPRINT-Twins compared to original SPRINT treatment and placebo arms. | 8 |
| Table S2: Baseline Characteristics of another 5 SPRINT-Twins compared to original SPRINT treatment and placebo arms. | 9 |
| Table S3: Accuracy or Mean Absolute Difference of Spearman Correlation between Variables for each RCT or Twin | 10 |
| Table S4: Baseline Characteristics of $SPRINT_{ACCORD}$ Twins compared to original SPRINT treatment and placebo arms. | 11 |
| Table S5: Baseline Characteristics of another 5 $SPRINT_{ACCORD}$ Twins compared to original SPRINT treatment and placebo arms. | 12 |
| Table S6: Baseline Characteristics of 5 $ACCORD_{SPRINT}$ Twins compared to original ACCORD treatment and placebo arms. | 13 |
| Table S7 Baseline Characteristics of another 5 $ACCORD_{SPRINT}$ Twins compared to original ACCORD treatment and placebo arms. | 14 |
| Table S8: Sensitivity Analysis of Training Sample Size and Model Parameters | 15-17 |
| Table S9: Statistical Comparison of Non-Conditioned Twin Generation Approaches-First Aggregate Level | 18 |
| Table S10: Statistical Comparison of Non-Conditioned Twin Generation Approaches-Second Aggregate Level | 19 |
| Table S11: Comparison of Machine Learning Efficiency of Non-Conditioned Twin Generation Approaches | 20 |
| Table S12: Baseline Characteristics of 5 $SPRINT_{EHR}$ Twins compared to original SPRINT treatment and placebo arms. | 21 |

| | |
|---|---|
| Table S13: Baseline Characteristics of another 5 $SPRINT_{EHR}$ Twins compared to original SPRINT treatment and placebo arms. | 22 |
| Table S14: Baseline Characteristics of 5 $ACCORD_{EHR}$ Twins compared to original ACCORD treatment and placebo arms. | 23 |
| Table S15: Baseline Characteristics of another 5 $ACCORD_{EHR}$ Twins compared to original ACCORD treatment and placebo arms. | 24 |
| Table S16: ICD-10-CDM codes used to extract EHR cohorts and covariates | 25-26 |
| Table S17: Covariates and bounds for continuous variables | 27 |
| Table S18: Summary of baseline features available from both SPRINT and ACCORD | 28-29 |
| Table S19: Pairs in Directed Acyclic Graph | 30-31 |
| Table S20: RCT-Twin-GAN Parameter search grid | 32 |

Figure S1: Spearman Correlations between variables within RCTs and Twins

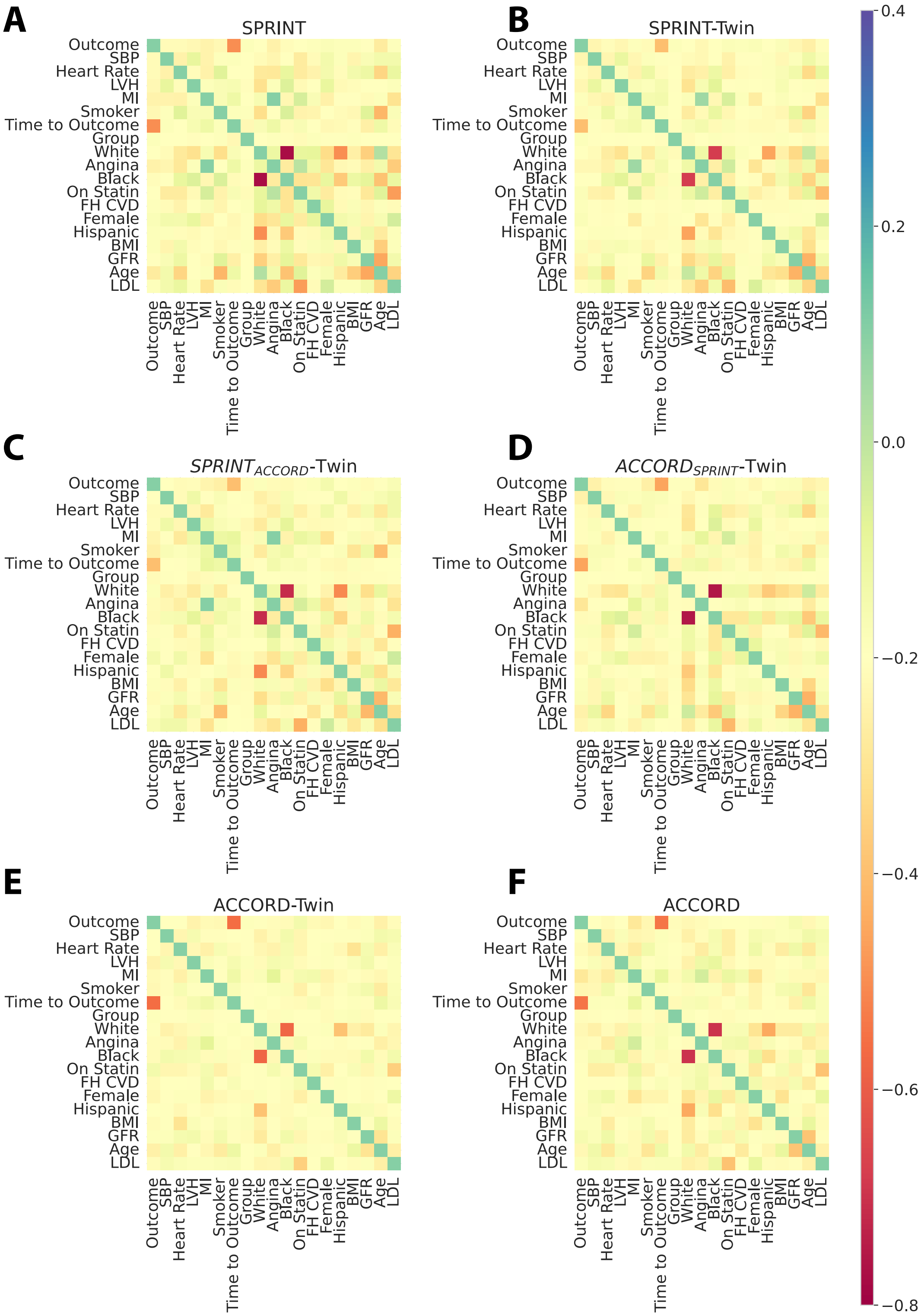

A-F is a heat map for a different RCT or digital twin. Abbreviations: FH: Family History, CVD: Cardiovascular disease, BMI: Body Mass Index, GFR: Glomerular Filtration Rate, LVH: Left ventricular hypertrophy, LDL: low-density lipoprotein, MI: Myocardial infarction, SBP: Systolic Blood Pressure.

Figure S2: Absolute standardized mean difference (ASMD) between RCTs and digital twins conditioned with the other RCT

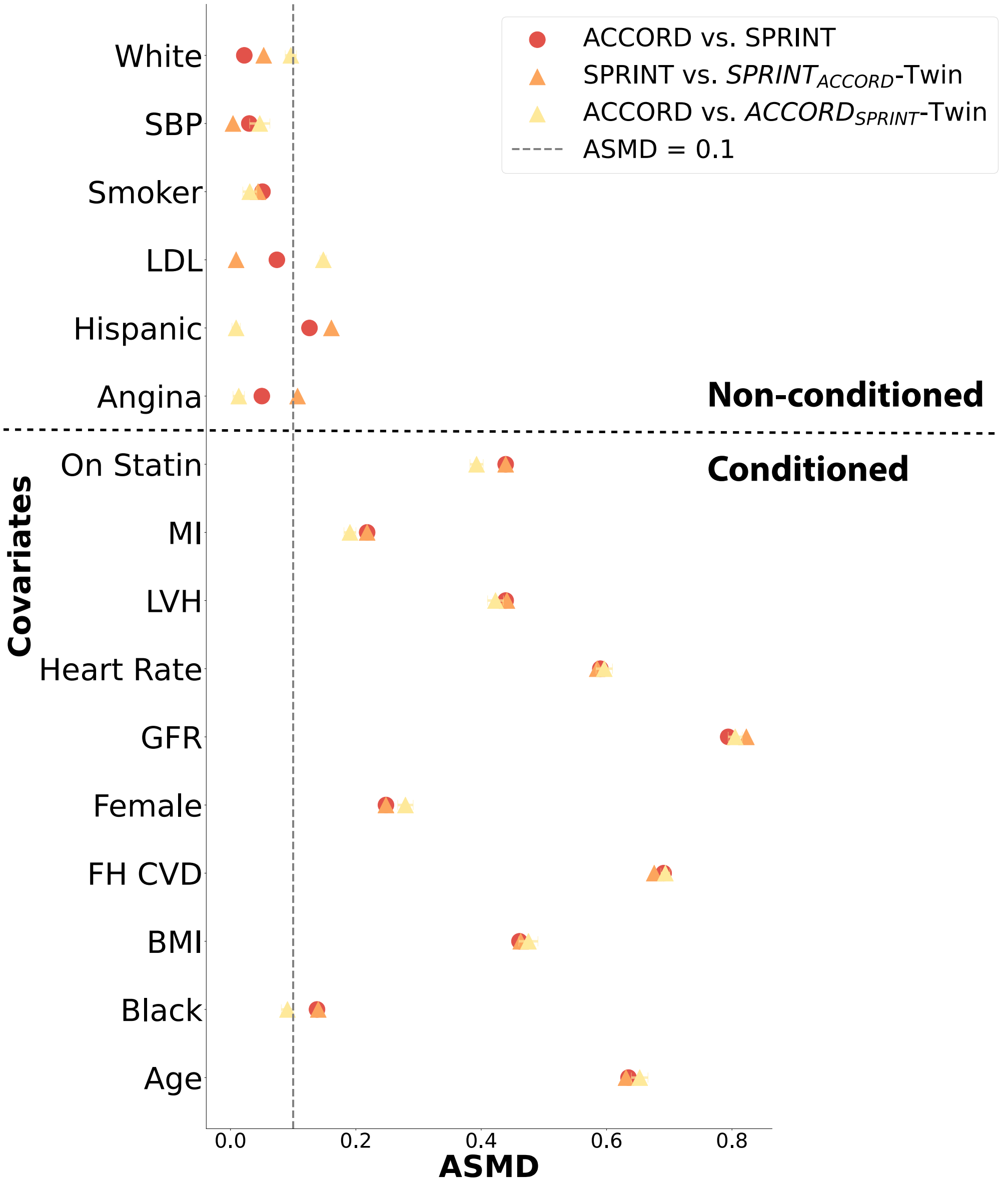

Markers include ACCORD vs. SPRINT (red circle), SPRINT vs $\text{SPRINT}_{\text{ACCORD}}$ Twin (orange triangle), and ACCORD vs $\text{ACCORD}_{\text{SPRINT}}$ Twin (yellow triangle). The grey dotted line represents an ASMD of 0.1, while the black dotted line separates the non-conditioned and conditioned covariates. Subscript is the conditioning cohort. The conditioning covariates included Age, Black, BMI, FH CVD, Female, GFR, Heart Rate, LVH, MI, and On Statin. Abbreviations: FH: Family History, CVD: Cardiovascular disease, BMI: Body Mass Index, GFR: Glomerular Filtration Rate, LVH: Left ventricular hypertrophy, LDL: low-density lipoprotein, MI: Myocardial infarction, SBP: Systolic Blood Pressure, SMD: Standardized Mean Difference, $\text{SPRINT}_{\text{ACCORD}}$-Twin: SPRINT conditioned on ACCORD digital twin, $\text{ACCORD}_{\text{SPRINT}}$Twin: ACCORD conditioned on SPRINT digital twin.

Figure S3: Single Variate Logistic Regression Accuracy of RCT vs Non-conditioned Twin

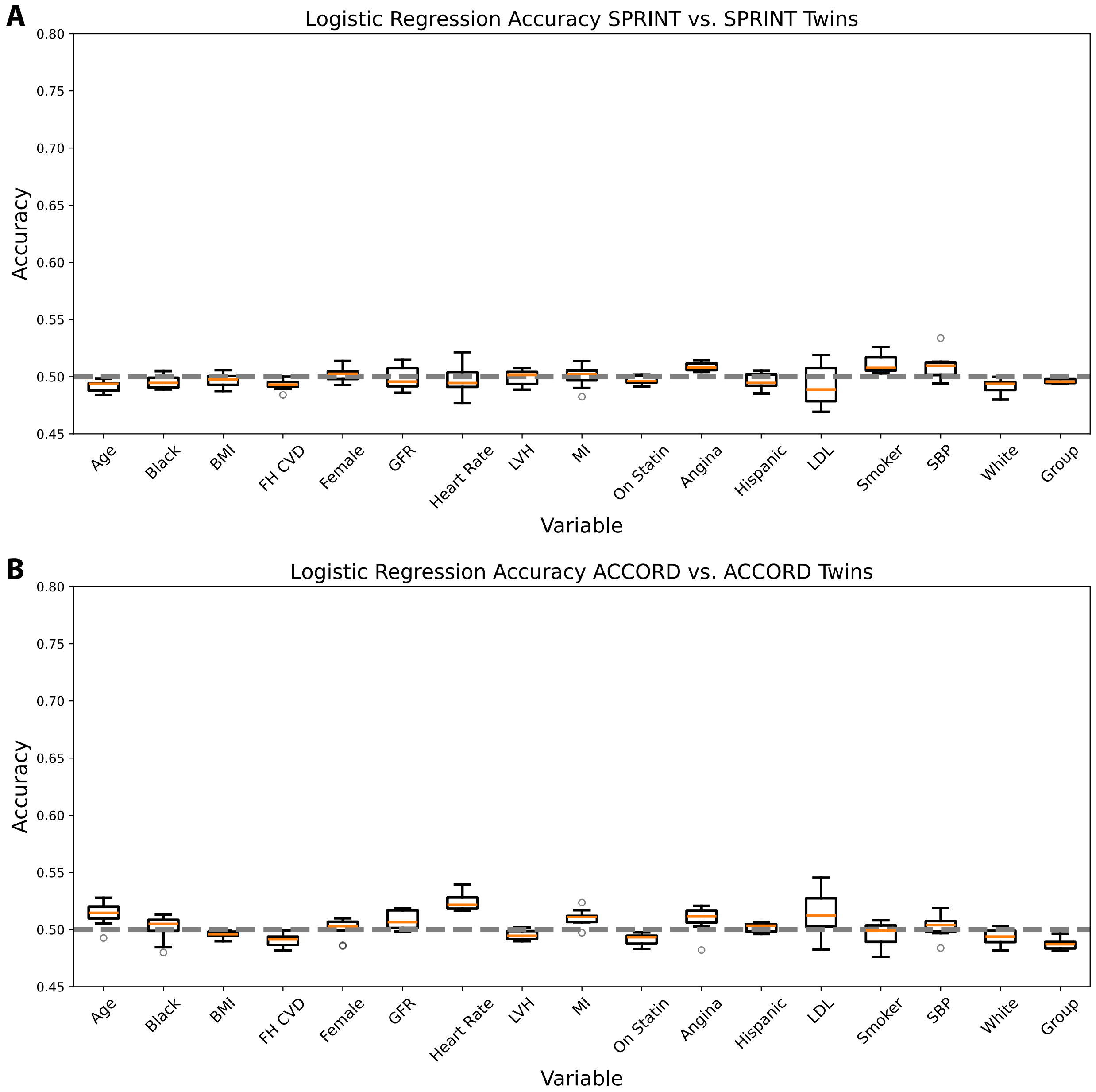

(A) Comparison of SPRINT vs. SPRINT twins, and (B) Comparison of ACCORD vs ACCORD twins. The gray dotted line represents statistical indistinguishability. Subscript is the conditioning cohort. Abbreviations: FH: Family History, CVD: Cardiovascular disease, BMI: Body Mass Index, GFR: Glomerular Filtration Rate, LVH: Left ventricular hypertrophy, LDL: low-density lipoprotein, MI: Myocardial infarction, SBP: Systolic Blood Pressure.

Figure S4: Absolute standardized mean difference (ASMD) between treatment and placebo arms of RCTs and EHR digital twins.

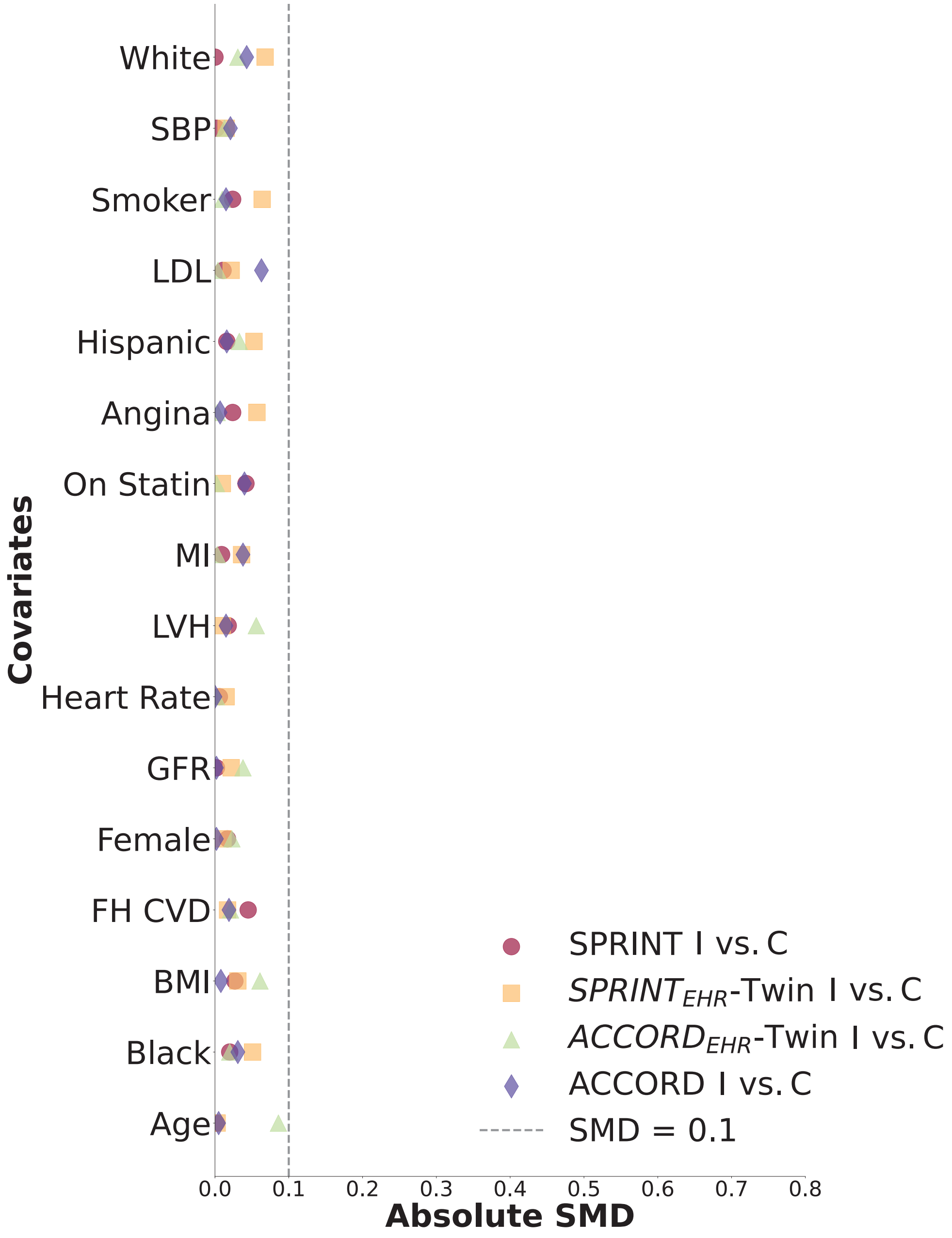

Markers include SPRINT (red circle), $SPRINT_{EHR}$-Twin (orange square), $ACCORD_{EHR}$ Twin (green triangle), and ACCORD (purple diamond The dotted line represents an ASMD of 0.1. Subscript is the conditioning cohort. The conditioning covariates included Age, Black, BMI, FH CVD, Female, GFR, Heart Rate, LVH, MI, and On Statin. Abbreviations: C: Control Arm, I: Intervention Arm, FH: Family History, CVD: Cardiovascular disease, BMI: Body Mass Index, GFR: Glomerular Filtration Rate, LVH: Left ventricular hypertrophy, LDL: low-density lipoprotein, MI: Myocardial infarction, SBP: Systolic Blood Pressure, SMD: Standardized Mean Difference, $SPRINT_{EHR}$ Twin: SPRINT conditioned on EHR digital twin, $ACCORD_{EHR}$ Twin: ACCORD conditioned on EHR digital twin

Figure S5: Representative Kaplan-Meier curves of digital twins conditioned on RCT data.

**A.**

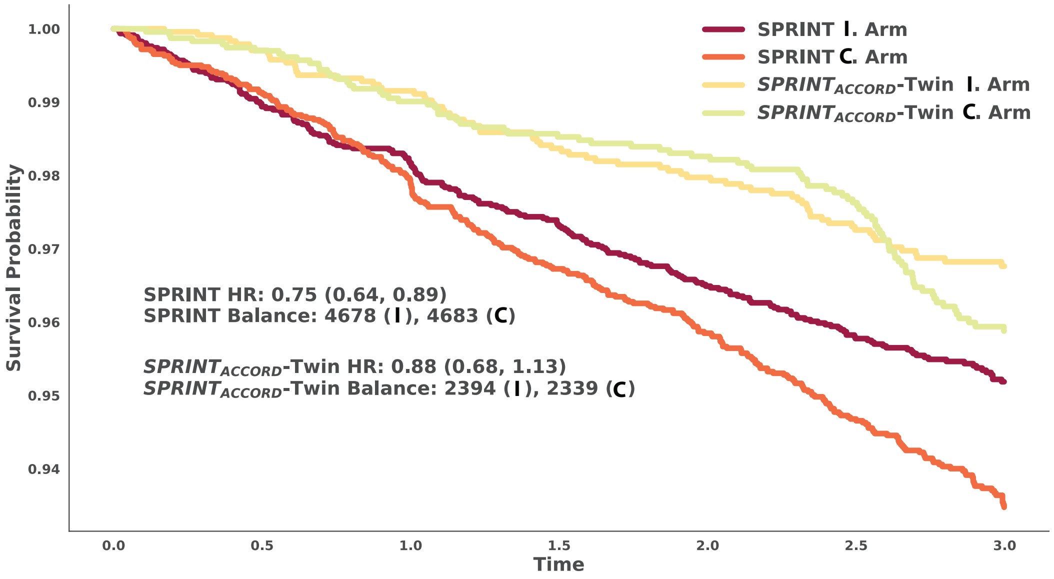

**B.**

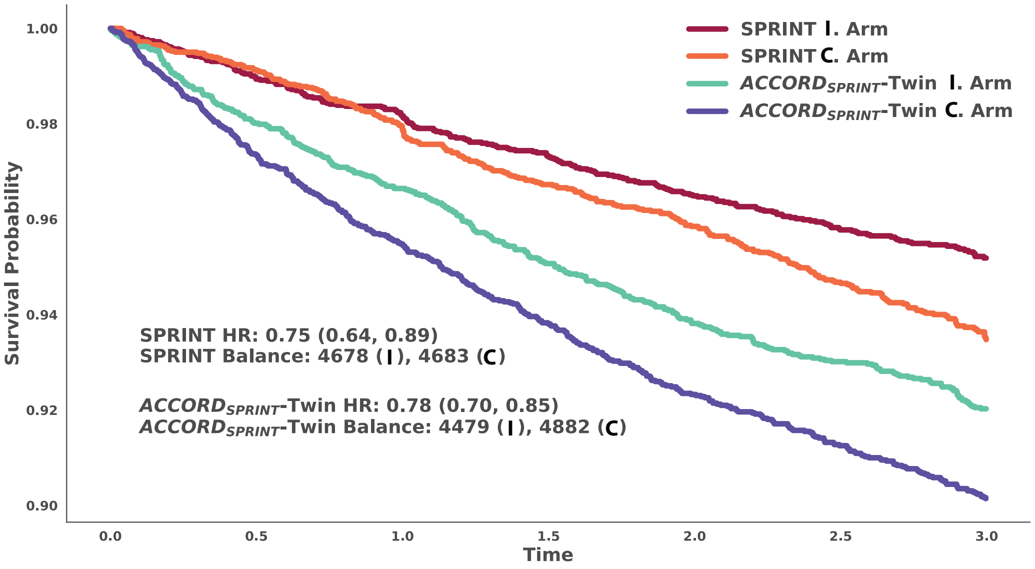

A) Kaplan-Meier curves of SPRINT treatment arms along with $\text{SPRINT}_{\text{ACCORD}}$ treatment arms, (B) Kaplan Meier curves of ACCORD treatment arms along with $\text{ACCORD}_{\text{SPRINT}}$ treatment arm balance of the original cohorts and digital twins. Abbreviations: C: Control Arm, I: Intervention Arm, $\text{SPRINT}_{\text{ACCORD}}$-Twin: SPRINT conditioned on ACCORD digital twin, $\text{ACCORD}_{\text{SPRINT}}$Twin: ACCORD conditioned on SPRINT digital twin.

Table S1: Baseline Characteristics of 5 SPRINT-Twins compared to original SPRINT treatment and placebo arms.

| Column | SPRINT C | SPRINT I | Twin 1 C | Twin 1 I | Twin 2 C | Twin 2 I | Twin 3 C | Twin 3 I | Twin 4 C | Twin 4 I | Twin 5 C | Twin 5 I |
|---|---|---|---|---|---|---|---|---|---|---|---|---|
| N | 4683 | 4678 | 4755 | 4606 | 4685 | 4676 | 4659 | 4702 | 4702 | 4659 | 4742 | 4619 |
| Age | 67[61,76] | 67[61,75] | 66[60,75] | 66[60,75] | 66[60,74] | 66[60,74] | 66[60,75] | 66[60,74] | 66[60,74] | 66[60,74] | 66[60,74] | 66[60,74] |
| Female | 1648(35) | 1684(35) | 1647(34) | 1552(33) | 1683(35) | 1627(34) | 1516(32) | 1704(36) | 1617(34) | 1562(33) | 1594(33) | 1559(33) |
| Black Race | 1423(30) | 1379(29) | 1415(29) | 1334(28) | 1369(29) | 1379(29) | 1327(28) | 1382(29) | 1391(29) | 1369(29) | 1367(28) | 1400(30) |
| Hispanic | 481(10) | 503(10) | 475(9) | 490(10) | 466(9) | 448(9) | 416(8) | 438(9) | 480(10) | 438(9) | 464(9) | 460(9) |
| White Race | 2701(57) | 2698(57) | 2779(58) | 2644(57) | 2751(58) | 2728(58) | 2780(59) | 2767(58) | 2721(57) | 2726(58) | 2834(59) | 2668(57) |
| BMI>30 | 1345(28) | 1402(29) | 1418(29) | 1329(28) | 1369(29) | 1329(28) | 1316(28) | 1291(27) | 1335(28) | 1269(27) | 1322(27) | 1310(28) |
| Systolic BP | 138[130,149] | 138[129,149] | 139[128,151] | 139[128,151] | 139[128,151] | 139[128,151] | 139[128,151] | 139[128,151] | 139[128,151] | 138[128,151] | 138[128,151] | 139[129,151] |
| Heart Rate | 65[58,74] | 65[58,73] | 65[57,75] | 65[57,75] | 66[57,75] | 66[57,75] | 65[57,75] | 65[57,75] | 66[57,76] | 65[57,75] | 66[57,75] | 65[57,75] |
| MI | 328(7) | 338(7) | 318(6) | 313(6) | 332(7) | 330(7) | 324(6) | 307(6) | 357(7) | 356(7) | 345(7) | 319(6) |
| LVH | 898(19) | 865(18) | 869(18) | 875(18) | 882(18) | 873(18) | 876(18) | 869(18) | 897(19) | 851(18) | 911(19) | 888(19) |
| On Statin | 2090(44) | 1990(42) | 1993(41) | 1931(41) | 2030(43) | 1991(42) | 2048(43) | 2014(42) | 2055(43) | 2003(42) | 2035(42) | 1935(41) |
| GFR | 71[58,84] | 71[58,84] | 71[57,85] | 72[57,85] | 72[57,85] | 72[57,85] | 72[57,85] | 72[57,85] | 72[57,85] | 72[57,85] | 72[58,86] | 72[58,86] |
| LDL | 110[88,134] | 110[87,133] | 110[85,138] | 109[85,136] | 109[84,137] | 111[86,137] | 109[84,137] | 110[86,138] | 110[86,138] | 110[85,138] | 109[85,136] | 110[85,137] |
| Smoker | 602(12) | 640(13) | 653(13) | 631(13) | 620(13) | 643(13) | 660(14) | 627(13) | 665(14) | 647(13) | 678(14) | 643(13) |
| Family Hist. CVD | 2965(63) | 3063(65) | 2986(62) | 2947(63) | 3032(64) | 3045(65) | 3033(65) | 3057(65) | 3057(65) | 3016(64) | 3036(64) | 2967(64) |
| Angina | 588(12) | 625(13) | 629(13) | 574(12) | 601(12) | 630(13) | 641(13) | 545(11) | 631(13) | 656(14) | 650(13) | 611(13) |

First column includes covariates, second and third column includes original SPRINT population characteristics, and subsequent columns include SPRINT-Twins. Abbreviations: C: Control Arm, I: Intervention Arm, FH: Family History, CVD: Cardiovascular disease, BMI: Body Mass Index, GFR: Glomerular Filtration Rate, LVH: Left ventricular hypertrophy, LDL: low-density lipoprotein, MI: Myocardial infarction, SBP: Systolic Blood Pressure

Table S2: Baseline Characteristics of another 5 SPRINT-Twins compared to original SPRINT treatment and placebo arms.

| Column | SPRINT C | SPRINT I | Twin 6 C | Twin 6 I | Twin 7 C | Twin 7 I | Twin 8 C | Twin 8 I | Twin 9 C | Twin 9 I | Twin 10 C | Twin 10 I |
|---|---|---|---|---|---|---|---|---|---|---|---|---|
| N | 4683 | 4678 | 4673 | 4688 | 4725 | 4636 | 4709 | 4652 | 4732 | 4629 | 4728 | 4633 |
| Age | 67[61,76] | 67[61,75] | 67[60,75] | 66[60,75] | 66[60,74] | 66[60,74] | 66[60,74] | 66[60,75] | 67[60,75] | 67[60,75] | 67[60,75] | 66[60,74] |
| Female | 1648(35) | 1684(35) | 1589(34) | 1608(34) | 1582(33) | 1624(35) | 1698(36) | 1665(35) | 1649(34) | 1613(34) | 1661(35) | 1623(35) |
| Black Race | 1423(30) | 1379(29) | 1337(28) | 1296(27) | 1344(28) | 1297(27) | 1420(30) | 1284(27) | 1320(27) | 1372(29) | 1312(27) | 1382(29) |
| Hispanic | 481(10) | 503(10) | 475(10) | 477(10) | 481(10) | 469(10) | 455(9) | 452(9) | 467(9) | 499(10) | 501(10) | 482(10) |
| White Race | 2701(57) | 2698(57) | 2750(58) | 2782(59) | 2779(58) | 2731(58) | 2748(58) | 2789(59) | 2872(60) | 2693(58) | 2813(59) | 2591(55) |
| BMI>30 | 1345(28) | 1402(29) | 1348(28) | 1364(29) | 1368(28) | 1336(28) | 1337(28) | 1339(28) | 1432(30) | 1288(27) | 1346(28) | 1346(29) |
| Systolic BP | 138[130,149] | 138[129,149] | 139[128,151] | 139[128,151] | 139[128,151] | 139[128,151] | 139[128,151] | 139[129,151] | 139[128,151] | 139[128,152] | 139[128,152] | 139[129,151] |
| Heart Rate | 65[58,74] | 65[58,73] | 65[57,75] | 65[57,75] | 66[57,75] | 66[57,75] | 65[57,75] | 66[57,75] | 66[57,75] | 66[57,75] | 65[57,75] | 65[57,75] |
| MI | 328(7) | 338(7) | 324(6) | 341(7) | 356(7) | 309(6) | 314(6) | 297(6) | 299(6) | 330(7) | 341(7) | 319(6) |
| LVH | 898(19) | 865(18) | 920(19) | 899(19) | 886(18) | 844(18) | 876(18) | 934(20) | 859(18) | 902(19) | 877(18) | 869(18) |
| On Statin | 2090(44) | 1990(42) | 2050(43) | 2012(42) | 2055(43) | 2004(43) | 2015(42) | 2003(43) | 2063(43) | 1946(42) | 2026(42) | 1953(42) |
| GFR | 71[58,84] | 71[58,84] | 72[57,85] | 72[57,85] | 72[57,85] | 72[57,86] | 72[57,86] | 72[57,85] | 71[57,85] | 72[57,86] | 72[58,85] | 72[57,85] |
| LDL | 110[88,134] | 110[87,133] | 110[85,138] | 109[84,137] | 109[85,136] | 109[85,137] | 110[86,137] | 109[85,136] | 109[85,137] | 109[85,137] | 109[85,137] | 109[84,137] |
| Smoker | 602(12) | 640(13) | 642(13) | 657(14) | 631(13) | 633(13) | 679(14) | 655(14) | 645(13) | 648(13) | 647(13) | 652(14) |
| Family Hist. CVD | 2965(63) | 3063(65) | 3078(65) | 3013(64) | 3039(64) | 2982(64) | 2992(63) | 2959(63) | 3055(64) | 2943(63) | 2994(63) | 3014(65) |
| Angina | 588(12) | 625(13) | 660(14) | 602(12) | 638(13) | 608(13) | 611(12) | 596(12) | 652(13) | 596(12) | 616(13) | 596(12) |

First column includes covariates, second and third column includes original SPRINT population characteristics, and subsequent columns include SPRINT-Twins. Abbreviations: C: Control Arm, I: Intervention Arm, FH: Family History, CVD: Cardiovascular disease, BMI: Body Mass Index, GFR: Glomerular Filtration Rate, LVH: Left ventricular hypertrophy, LDL: low-density lipoprotein, MI: Myocardial infarction, SBP: Systolic Blood Pressure

Table S3: Accuracy or Mean Absolute Difference of Spearman Correlation between Variables for each RCT or Twin

| **Model** | SPRINT-Twin | | $SPRINT_{ACCORD}$ Twin | | $ACCORD_{SPRINT}$ Twin | | ACCORD Twin | | ACCORD | |
|---|---|---|---|---|---|---|---|---|---|---|
| **Measure** | CorAcc (%) | MAD | CorAcc (%) | MAD | CorAcc (%) | MAD | CorAcc (%) | MAD | CorAcc (%) | MAD |
| SPRINT | **88.37%** | **0.03** | 78.39% | 0.05 | **85.60%** | **0.04** | 72.85% | 0.06 | 77.84% | 0.05 |
| SPRINT Twin | | | **85.60%** | **0.04** | 83.93% | 0.04 | 82.27% | 0.04 | 82.83% | 0.04 |
| $SPRINT_{ACCORD}$ Twin | | | | | 76.73% | 0.05 | 80.61% | 0.05 | 85.60% | 0.04 |
| $ACCORD_{SPRINT}$ Twin | | | | | | | 82.83% | 0.05 | 84.49% | 0.04 |
| ACCORD Twin | | | | | | | | | **89.47%** | **0.03** |

Accuracy of Spearman Correlation measurement or MAD in column cohorts compared to row cohorts. Bold is the highest accuracy and lowest MAD for each cohort. Abbreviations: CorAcc: Accuracy of Spearman Correlation in Column cohort compared to row cohort, MAD: Mean Absolute Difference, $SPRINT_{ACCORD}$-Twin: SPRINT conditioned on ACCORD digital twin, $ACCORD_{SPRINT}$Twin: ACCORD conditioned on SPRINT digital twin

Table S4: Baseline Characteristics of $\text{SPRINT}_{\text{ACCORD}}$ Twins compared to original SPRINT treatment and placebo arms.

| Column | SPRINT C | SPRINT I | Twin 1 C | Twin 1 I | Twin 2 C | Twin 2 I | Twin 3 C | Twin 3 I | Twin 4 C | Twin 4 I | Twin 5 C | Twin 5 I |
|---|---|---|---|---|---|---|---|---|---|---|---|---|
| N | 4683 | 4678 | 2347 | 2386 | 2327 | 2406 | 2339 | 2394 | 2340 | 2393 | 2382 | 2351 |
| Age | 67[61,76] | 67[61,75] | 62[58,67] | 62[58,67] | 62[57,67] | 62[58,67] | 62[58,67] | 62[58,67] | 62[58,67] | 62[58,67] | 62[58,68] | 62[58,67] |
| Female | 1648(35) | 1684(35) | 1081(46) | 1160(48) | 1106(47) | 1141(47) | 1100(47) | 1160(48) | 1124(48) | 1153(48) | 1119(46) | 1121(47) |
| Black Race | 1423(30) | 1379(29) | 553(23) | 556(23) | 576(24) | 594(24) | 546(23) | 578(24) | 535(22) | 584(24) | 550(23) | 587(24) |
| Hispanic | 481(10) | 503(10) | 402(17) | 373(15) | 414(17) | 407(16) | 393(16) | 362(15) | 438(18) | 387(16) | 374(15) | 385(16) |
| White Race | 2701(57) | 2698(57) | 1402(59) | 1442(60) | 1319(56) | 1407(58) | 1407(60) | 1446(60) | 1353(57) | 1420(59) | 1453(60) | 1374(58) |
| BMI>30 | 1345(28) | 1402(29) | 1226(52) | 1250(52) | 1202(51) | 1236(51) | 1220(52) | 1216(50) | 1198(51) | 1243(51) | 1286(53) | 1196(50) |
| Systolic BP | 138[130,149] | 138[129,149] | 138[129,149] | 139[130,150] | 139[130,149] | 139[130,149] | 138[129,149] | 139[129,149] | 139[130,149] | 139[130,149] | 138[129,148] | 139[130,149] |
| Heart Rate | 65[58,74] | 65[58,73] | 72[65,80] | 72[65,80] | 73[65,81] | 72[65,80] | 72[65,81] | 72[65,80] | 72[65,80] | 72[65,81] | 72[65,81] | 73[65,81] |
| MI | 328(7) | 338(7) | 334(14) | 316(13) | 320(13) | 341(14) | 342(14) | 309(12) | 321(13) | 331(13) | 357(14) | 336(14) |
| LVH | 898(19) | 865(18) | 99(4) | 114(4) | 129(5) | 124(5) | 131(5) | 101(4) | 101(4) | 111(4) | 125(5) | 123(5) |
| On Statin | 2090(44) | 1990(42) | 1527(65) | 1553(65) | 1492(64) | 1598(66) | 1508(64) | 1565(65) | 1535(65) | 1558(65) | 1560(65) | 1498(63) |
| GFR | 71[58,84] | 71[58,84] | 89[75,105] | 89[73,104] | 89[75,105] | 89[75,105] | 89[75,105] | 89[74,105] | 90[76,105] | 89[75,105] | 89[75,105] | 89[76,105] |
| LDL | 110[88,134] | 110[87,133] | 109[82,135] | 109[86,137] | 108[82,136] | 107[82,136] | 109[83,137] | 108[84,136] | 107[85,135] | 109[82,135] | 106[81,135] | 109[85,135] |
| Smoker | 602(12) | 640(13) | 343(14) | 372(15) | 388(16) | 398(16) | 332(14) | 368(15) | 354(15) | 382(15) | 352(14) | 317(13) |
| Family Hist. CVD | 2965(63) | 3063(65) | 722(30) | 751(31) | 699(30) | 722(30) | 759(32) | 774(32) | 711(30) | 780(32) | 761(31) | 731(31) |
| Angina | 588(12) | 625(13) | 445(18) | 413(17) | 415(17) | 426(17) | 387(16) | 406(16) | 379(16) | 401(16) | 425(17) | 401(17) |

First column includes covariates, second and third column includes original SPRINT population characteristics, and subsequent columns include $\text{SPRINT}_{\text{ACCORD}}$-Twins. Abbreviations: C: Control Arm, I: Intervention Arm, FH: Family History, CVD: Cardiovascular disease, BMI: Body Mass Index, GFR: Glomerular Filtration Rate, LVH: Left ventricular hypertrophy, LDL: low-density lipoprotein, MI: Myocardial infarction, SBP: Systolic Blood Pressure

Table S5: Baseline Characteristics of another 5 $\text{SPRINT}_{\text{ACCORD}}$ Twins compared to original SPRINT treatment and placebo arms.

| Column | SPRINT C | SPRINT I | Twin 1 C | Twin 1 I | Twin 2 C | Twin 2 I | Twin 3 C | Twin 3 I | Twin 4 C | Twin 4 I | Twin 5 C | Twin 5 I |
|---|---|---|---|---|---|---|---|---|---|---|---|---|
| N | 4683 | 4678 | 2361 | 2372 | 2295 | 2438 | 2352 | 2381 | 2372 | 2361 | 2336 | 2397 |
| Age | 67[61,76] | 67[61,75] | 62[58,67] | 62[57,67] | 62[58,67] | 62[58,67] | 62[58,67] | 62[58,67] | 62[58,67] | 62[58,67] | 62[58,68] | 62[58,67] |
| Female | 1648(35) | 1684(35) | 1133(47) | 1178(49) | 1114(48) | 1175(48) | 1154(49) | 1131(47) | 1122(47) | 1123(47) | 1153(49) | 1106(46) |
| Black Race | 1423(30) | 1379(29) | 567(24) | 549(23) | 550(23) | 582(23) | 542(23) | 571(23) | 602(25) | 576(24) | 563(24) | 559(23) |
| Hispanic | 481(10) | 503(10) | 370(15) | 407(17) | 360(15) | 378(15) | 397(16) | 380(15) | 382(16) | 390(16) | 400(17) | 427(17) |
| White Race | 2701(57) | 2698(57) | 1419(60) | 1402(59) | 1380(60) | 1469(60) | 1396(59) | 1414(59) | 1371(57) | 1358(57) | 1353(57) | 1411(58) |
| BMI>30 | 1345(28) | 1402(29) | 1193(50) | 1192(50) | 1176(51) | 1233(50) | 1209(51) | 1182(49) | 1225(51) | 1196(50) | 1183(50) | 1231(51) |
| Systolic BP | 138[130,149] | 138[129,149] | 138[129,148] | 139[129,150] | 139[130,149] | 140[131,150] | 139[131,150] | 138[129,149] | 138[129,149] | 139[130,149] | 139[130,150] | 139[130,149] |
| Heart Rate | 65[58,74] | 65[58,73] | 73[66,80] | 72[65,81] | 72[65,80] | 73[65,81] | 72[65,80] | 73[65,81] | 72[65,80] | 72[65,81] | 73[65,81] | 72[65,80] |
| MI | 328(7) | 338(7) | 323(13) | 327(13) | 313(13) | 360(14) | 327(13) | 370(15) | 326(13) | 327(13) | 305(13) | 361(15) |
| LVH | 898(19) | 865(18) | 117(4) | 115(4) | 98(4) | 134(5) | 125(5) | 131(5) | 126(5) | 121(5) | 106(4) | 130(5) |
| On Statin | 2090(44) | 1990(42) | 1480(62) | 1522(64) | 1505(65) | 1601(65) | 1538(65) | 1541(64) | 1577(66) | 1486(62) | 1524(65) | 1537(64) |
| GFR | 71[58,84] | 71[58,84] | 90[76,105] | 89[76,105] | 89[76,105] | 89[75,105] | 89[76,106] | 89[75,105] | 89[75,105] | 90[76,106] | 89[74,105] | 89[76,105] |
| LDL | 110[88,134] | 110[87,133] | 109[85,136] | 108[84,135] | 109[85,137] | 109[85,135] | 107[83,133] | 108[84,136] | 108[83,135] | 110[85,138] | 107[83,136] | 108[85,137] |
| Smoker | 602(12) | 640(13) | 399(16) | 374(15) | 342(14) | 420(17) | 364(15) | 341(14) | 379(15) | 368(15) | 360(15) | 378(15) |
| Family Hist. CVD | 2965(63) | 3063(65) | 771(32) | 763(32) | 745(32) | 776(31) | 733(31) | 753(31) | 750(31) | 771(32) | 762(32) | 740(30) |
| Angina | 588(12) | 625(13) | 408(17) | 387(16) | 395(17) | 450(18) | 398(16) | 446(18) | 403(16) | 405(17) | 415(17) | 434(18) |

First column includes covariates, second and third column includes original SPRINT population characteristics, and subsequent columns include $\text{SPRINT}_{\text{ACCORD}}$-Twins. Abbreviations: C: Control Arm, I: Intervention Arm, FH: Family History, CVD: Cardiovascular disease, BMI: Body Mass Index, GFR: Glomerular Filtration Rate, LVH: Left ventricular hypertrophy, LDL: low-density lipoprotein, MI: Myocardial infarction, SBP: Systolic Blood Pressure

Table S6: Baseline Characteristics of 5 ACCORD$_{SPRINT}$ Twins compared to original ACCORD treatment and placebo arms.

| Column | ACCORD C | ACCORD I | Twin 1 C | Twin 1 I | Twin 2 C | Twin 2 I | Twin 3 C | Twin 3 I | Twin 4 C | Twin 4 I | Twin 5 C | Twin 5 I |
|---|---|---|---|---|---|---|---|---|---|---|---|---|
| N | 2371 | 2362 | 4749 | 4612 | 4751 | 4610 | 4776 | 4585 | 4756 | 4605 | 4746 | 4615 |
| Age | 62[57,67] | 61[57,67] | 67[61,76] | 67[61,75] | 67[61,76] | 68[61,76] | 67[61,76] | 67[61,76] | 67[61,75] | 67[61,76] | 67[61,76] | 68[61,76] |
| Female | 1130(47) | 1128(47) | 1622(34) | 1568(33) | 1658(34) | 1594(34) | 1585(33) | 1532(33) | 1698(35) | 1578(34) | 1612(33) | 1608(34) |
| Black Race | 580(24) | 547(23) | 1276(26) | 1274(27) | 1316(27) | 1298(28) | 1384(28) | 1274(27) | 1323(27) | 1326(28) | 1298(27) | 1239(26) |
| Hispanic | 170(7) | 160(6) | 293(6) | 313(6) | 296(6) | 338(7) | 315(6) | 322(7) | 303(6) | 306(6) | 324(6) | 311(6) |
| White Race | 1368(57) | 1413(59) | 3050(64) | 2922(63) | 3052(64) | 2883(62) | 3003(62) | 2912(63) | 3048(64) | 2878(62) | 3011(63) | 2961(64) |
| BMI>30 | 1213(51) | 1218(51) | 1365(28) | 1365(29) | 1364(28) | 1271(27) | 1378(28) | 1302(28) | 1373(28) | 1349(29) | 1365(28) | 1347(29) |
| Systolic BP | 138[130,149] | 138[128,149] | 139[125,152] | 138[124,151] | 138[125,151] | 139[125,152] | 139[125,152] | 139[125,152] | 138[125,150] | 139[125,152] | 139[125,152] | 139[125,152] |
| Heart Rate | 72[65,81] | 72[65,80] | 65[58,73] | 65[58,73] | 65[58,74] | 65[58,74] | 65[58,73] | 65[58,73] | 65[58,74] | 65[58,73] | 65[58,74] | 65[58,74] |
| MI | 341(14) | 309(13) | 365(7) | 343(7) | 384(8) | 349(7) | 399(8) | 387(8) | 364(7) | 385(8) | 381(8) | 382(8) |
| LVH | 121(5) | 113(4) | 856(18) | 848(18) | 824(17) | 876(19) | 840(17) | 783(17) | 885(18) | 886(19) | 866(18) | 825(17) |
| On Statin | 1562(65) | 1511(63) | 2163(45) | 2053(44) | 2192(46) | 2148(46) | 2216(46) | 2105(45) | 2152(45) | 2139(46) | 2109(44) | 2124(46) |
| GFR | 89[75,105] | 89[75,105] | 71[57,84] | 71[58,84] | 71[58,84] | 71[57,84] | 71[57,84] | 71[58,84] | 71[57,84] | 70[57,84] | 71[58,84] | 70[56,83] |
| LDL | 105[85,129] | 106[86,133] | 100[73,130] | 101[74,133] | 101[73,132] | 101[73,130] | 101[73,132] | 100[73,130] | 100[73,130] | 100[73,131] | 100[73,131] | 101[73,132] |
| Smoker | 269(11) | 279(11) | 485(10) | 507(10) | 504(10) | 504(10) | 527(11) | 486(10) | 524(11) | 516(11) | 509(10) | 483(10) |
| Family Hist. CVD | 743(31) | 761(32) | 3061(64) | 2962(64) | 3040(63) | 3006(65) | 3045(63) | 2982(65) | 3116(65) | 2975(64) | 3021(63) | 2988(64) |
| Angina | 266(11) | 270(11) | 524(11) | 492(10) | 524(11) | 498(10) | 530(11) | 519(11) | 515(10) | 506(10) | 549(11) | 518(11) |

First column includes covariates, second and third column includes original ACCORD population characteristics, and subsequent columns include ACCORD$_{SPRINT}$-Twins. Abbreviations: C: Control Arm, I: Intervention Arm, FH: Family History, CVD: Cardiovascular disease, BMI: Body Mass Index, GFR: Glomerular Filtration Rate, LVH: Left ventricular hypertrophy, LDL: low-density lipoprotein, MI: Myocardial infarction, SBP: Systolic Blood Pressure

Table S7: Baseline Characteristics of another 5 $ACCORD_{SPRINT}$ Twins compared to original ACCORD treatment and placebo arms.

| Column | ACCORD C | ACCORD I | Twin 1 C | Twin 1 I | Twin 2 C | Twin 2 I | Twin 3 C | Twin 3 I | Twin 4 C | Twin 4 I | Twin 5 C | Twin 5 I |
|---|---|---|---|---|---|---|---|---|---|---|---|---|
| N | 2371 | 2362 | 4701 | 4660 | 4882 | 4479 | 4800 | 4561 | 4642 | 4719 | 4778 | 4583 |
| Age | 62[57,67] | 61[57,67] | 68[61,76] | 68[61,76] | 67[61,76] | 67[61,76] | 67[61,76] | 68[61,76] | 67[61,76] | 67[61,76] | 67[61,76] | 67[61,75] |
| Female | 1130(47) | 1128(47) | 1610(34) | 1625(34) | 1677(34) | 1510(33) | 1659(34) | 1531(33) | 1571(33) | 1545(32) | 1594(33) | 1564(34) |
| Black Race | 580(24) | 547(23) | 1297(27) | 1323(28) | 1391(28) | 1210(27) | 1348(28) | 1202(26) | 1303(28) | 1327(28) | 1310(27) | 1293(28) |
| Hispanic | 170(7) | 160(6) | 341(7) | 319(6) | 332(6) | 322(7) | 348(7) | 337(7) | 318(6) | 329(6) | 340(7) | 288(6) |
| White Race | 1368(57) | 1413(59) | 2970(63) | 2922(62) | 3081(63) | 2885(64) | 3015(62) | 2957(64) | 2914(62) | 2945(62) | 3070(64) | 2909(63) |
| BMI>30 | 1213(51) | 1218(51) | 1247(26) | 1320(28) | 1441(29) | 1221(27) | 1402(29) | 1326(29) | 1416(30) | 1371(29) | 1333(27) | 1320(28) |
| Systolic BP | 138[130,149] | 138[128,149] | 138[125,152] | 139[125,151] | 138[125,151] | 139[126,152] | 138[125,151] | 138[124,151] | 138[125,151] | 138[124,150] | 138[125,151] | 138[124,150] |
| Heart Rate | 72[65,81] | 72[65,80] | 65[58,73] | 65[58,74] | 65[58,74] | 65[58,73] | 65[58,74] | 64[57,73] | 65[58,73] | 65[58,73] | 65[59,74] | 65[58,74] |
| MI | 341(14) | 309(13) | 351(7) | 371(7) | 387(7) | 324(7) | 377(7) | 339(7) | 351(7) | 386(8) | 369(7) | 356(7) |
| LVH | 121(5) | 113(4) | 823(17) | 843(18) | 932(19) | 837(18) | 912(19) | 819(17) | 873(18) | 829(17) | 835(17) | 819(17) |
| On Statin | 1562(65) | 1511(63) | 2176(46) | 2148(46) | 2198(45) | 2038(45) | 2179(45) | 2069(45) | 2155(46) | 2173(46) | 2220(46) | 2094(45) |
| GFR | 89[75,105] | 89[75,105] | 70[57,83] | 70[57,83] | 71[58,84] | 71[57,84] | 71[58,85] | 70[57,82] | 71[58,84] | 71[58,84] | 71[58,84] | 71[58,84] |
| LDL | 105[85,129] | 106[86,133] | 100[73,131] | 101[73,131] | 101[73,133] | 100[72,130] | 101[74,132] | 101[72,133] | 101[73,131] | 101[73,132] | 100[73,131] | 100[73,132] |
| Smoker | 269(11) | 279(11) | 454(9) | 481(10) | 526(10) | 453(10) | 537(11) | 498(10) | 512(11) | 458(9) | 471(9) | 500(10) |
| Family Hist. CVD | 743(31) | 761(32) | 3053(64) | 3036(65) | 3136(64) | 2880(64) | 3083(64) | 2965(65) | 3027(65) | 3012(63) | 3045(63) | 2978(64) |
| Angina | 266(11) | 270(11) | 581(12) | 501(10) | 553(11) | 458(10) | 518(10) | 472(10) | 512(11) | 531(11) | 510(10) | 459(10) |

First column includes covariates, second and third column includes original ACCORD population characteristics, and subsequent columns include $ACCORD_{SPRINT}$-Twins. Abbreviations: C: Control Arm, I: Intervention Arm, FH: Family History, CVD: Cardiovascular disease, BMI: Body Mass Index, GFR: Glomerular Filtration Rate, LVH: Left ventricular hypertrophy, LDL: low-density lipoprotein, MI: Myocardial infarction, SBP: Systolic Blood Pressure

Table S8: Sensitivity Analysis of Training Sample Size and Model Parameters

| Pct. Sample Size | Batch Size | No. of Epochs | Mean HR | Lower bound 95% CI | Upper bound 95% CI | Pct. SC Arm | Pct. IC Arm | Converged? |
|---|---|---|---|---|---|---|---|---|
| 1 | 200 | 200 | 0.69 | 0.54 | 0.89 | 50 | 50 | 1 |
| 1 | 200 | 300 | 0.66 | 0.5 | 0.88 | 49.6 | 50.4 | 0 |
| 1 | 200 | 400 | 0.63 | 0.49 | 0.82 | 49.6 | 50.4 | 1 |
| 1 | 200 | 500 | 0.73 | 0.57 | 0.94 | 50.4 | 49.6 | 0 |
| 1 | 300 | 200 | 0.57 | 0.43 | 0.74 | 50 | 50 | 1 |
| 1 | 300 | 300 | 1.1 | 0.85 | 1.42 | 49.9 | 50.1 | 1 |
| 1 | 300 | 400 | 0.84 | 0.62 | 1.14 | 52.3 | 47.7 | 1 |
| 1 | 300 | 500 | 0.75 | 0.56 | 0.99 | 49.8 | 50.2 | 1 |
| 1 | 400 | 200 | 0.64 | 0.48 | 0.85 | 50.8 | 49.2 | 1 |
| 1 | 400 | 300 | 0.94 | 0.73 | 1.22 | 50.9 | 49.1 | 1 |
| 1 | 400 | 400 | 0.36 | 0.26 | 0.48 | 50.6 | 49.4 | 1 |
| 1 | 400 | 500 | 0.7 | 0.53 | 0.93 | 50.8 | 49.2 | 1 |
| 1 | 500 | 200 | 0.54 | 0.41 | 0.72 | 50 | 50 | 0 |
| 1 | 500 | 300 | 0.78 | 0.58 | 1.04 | 49.8 | 50.2 | 1 |
| 1 | 500 | 400 | 0.73 | 0.55 | 0.95 | 50 | 50 | 1 |
| 1 | 500 | 500 | 0.74 | 0.57 | 0.96 | 48.9 | 51.1 | 1 |
| 5 | 200 | 200 | 0.84 | 0.65 | 1.08 | 50.1 | 49.9 | 1 |
| 5 | 200 | 300 | 0.9 | 0.7 | 1.15 | 49.1 | 50.9 | 0 |
| 5 | 200 | 400 | 0.86 | 0.65 | 1.13 | 49.1 | 50.9 | 1 |
| 5 | 200 | 500 | 0.76 | 0.59 | 0.99 | 49.5 | 50.5 | 0 |
| 5 | 300 | 200 | 0.77 | 0.6 | 0.98 | 48.8 | 51.2 | 1 |
| 5 | 300 | 300 | 0.74 | 0.56 | 0.96 | 49.3 | 50.7 | 1 |
| 5 | 300 | 400 | 1.16 | 0.87 | 1.55 | 51 | 49 | 1 |
| 5 | 300 | 500 | 0.61 | 0.45 | 0.82 | 50.1 | 49.9 | 1 |
| 5 | 400 | 200 | 0.57 | 0.42 | 0.76 | 51 | 49 | 1 |
| 5 | 400 | 300 | 0.69 | 0.53 | 0.91 | 48.5 | 51.5 | 1 |
| 5 | 400 | 400 | 0.81 | 0.62 | 1.05 | 48.7 | 51.3 | 1 |
| 5 | 400 | 500 | 0.72 | 0.55 | 0.94 | 51.1 | 48.9 | 1 |
| 5 | 500 | 200 | 0.59 | 0.46 | 0.75 | 49 | 51 | 1 |
| 5 | 500 | 300 | 0.72 | 0.54 | 0.95 | 50 | 50 | 1 |
| 5 | 500 | 400 | 0.66 | 0.5 | 0.86 | 49.2 | 50.8 | 1 |
| 5 | 500 | 500 | 0.77 | 0.6 | 0.98 | 49.5 | 50.5 | 1 |
| 10 | 200 | 200 | 0.68 | 0.47 | 0.98 | 49.6 | 50.4 | 0 |
| 10 | 200 | 300 | 0.82 | 0.62 | 1.08 | 53 | 47 | 1 |
| 10 | 200 | 400 | 0.82 | 0.64 | 1.06 | 50.7 | 49.3 | 0 |

| | | | | | | | | |
|---|---|---|---|---|---|---|---|---|
| 10 | 200 | 500 | 0.58 | 0.42 | 0.79 | 49.5 | 50.5 | 1 |
| 10 | 300 | 200 | 0.91 | 0.72 | 1.14 | 50.3 | 49.7 | 1 |
| 10 | 300 | 300 | 0.91 | 0.7 | 1.18 | 49.2 | 50.8 | 1 |
| 10 | 300 | 400 | 0.74 | 0.54 | 1.01 | 48.4 | 51.6 | 1 |
| 10 | 300 | 500 | 0.92 | 0.7 | 1.19 | 50.2 | 49.8 | 1 |
| 10 | 400 | 200 | 0.42 | 0.31 | 0.56 | 49.6 | 50.4 | 1 |
| 10 | 400 | 300 | 0.77 | 0.57 | 1.04 | 52.1 | 47.9 | 1 |
| 10 | 400 | 400 | 0.66 | 0.51 | 0.85 | 49.8 | 50.2 | 1 |
| 10 | 400 | 500 | 0.94 | 0.72 | 1.24 | 50.8 | 49.2 | 1 |
| 10 | 500 | 200 | 0.54 | 0.42 | 0.7 | 49.7 | 50.3 | 1 |
| 10 | 500 | 300 | 0.49 | 0.36 | 0.67 | 50.6 | 49.4 | 1 |
| 10 | 500 | 400 | 0.63 | 0.49 | 0.81 | 50.4 | 49.6 | 1 |
| 10 | 500 | 500 | 0.61 | 0.47 | 0.8 | 50.2 | 49.8 | 1 |
| 25 | 200 | 200 | 1.11 | 0.86 | 1.44 | 51 | 49 | 1 |
| 25 | 200 | 300 | 0.67 | 0.52 | 0.87 | 50 | 50 | 1 |
| 25 | 200 | 400 | 0.64 | 0.47 | 0.87 | 49.6 | 50.4 | 0 |
| 25 | 200 | 500 | 0.77 | 0.58 | 1.03 | 49.3 | 50.7 | 1 |
| 25 | 300 | 200 | 0.49 | 0.35 | 0.69 | 50 | 50 | 1 |
| 25 | 300 | 300 | 0.72 | 0.54 | 0.96 | 51.1 | 48.9 | 0 |
| 25 | 300 | 400 | 0.85 | 0.64 | 1.12 | 52.5 | 47.5 | 0 |
| 25 | 300 | 500 | 0.74 | 0.58 | 0.96 | 48.6 | 51.4 | 1 |
| 25 | 400 | 200 | 0.81 | 0.59 | 1.12 | 50.4 | 49.6 | 1 |
| 25 | 400 | 300 | 0.8 | 0.59 | 1.08 | 50 | 50 | 1 |
| 25 | 400 | 400 | 0.53 | 0.4 | 0.7 | 50.3 | 49.7 | 1 |
| 25 | 400 | 500 | 0.71 | 0.53 | 0.93 | 49.1 | 50.9 | 1 |
| 25 | 500 | 200 | 0.62 | 0.44 | 0.87 | 50.6 | 49.4 | 1 |
| 25 | 500 | 300 | 0.71 | 0.54 | 0.95 | 50.6 | 49.4 | 1 |
| 25 | 500 | 400 | 0.9 | 0.66 | 1.24 | 48.9 | 51.1 | 1 |
| 25 | 500 | 500 | 0.49 | 0.37 | 0.64 | 49 | 51 | 1 |
| 50 | 200 | 200 | 0.88 | 0.69 | 1.12 | 49.5 | 50.5 | 1 |
| 50 | 200 | 300 | 0.87 | 0.65 | 1.17 | 49.7 | 50.3 | 1 |
| 50 | 200 | 400 | 0.8 | 0.61 | 1.03 | 49.3 | 50.7 | 0 |
| 50 | 200 | 500 | 0.82 | 0.64 | 1.06 | 51.2 | 48.8 | 1 |
| 50 | 300 | 200 | 0.64 | 0.49 | 0.83 | 49.6 | 50.4 | 1 |
| 50 | 300 | 300 | 0.84 | 0.65 | 1.1 | 50.4 | 49.6 | 1 |
| 50 | 300 | 400 | 0.8 | 0.61 | 1.07 | 50.7 | 49.3 | 1 |
| 50 | 300 | 500 | 0.77 | 0.58 | 1.02 | 50 | 50 | 1 |
| 50 | 400 | 200 | 0.69 | 0.54 | 0.9 | 49.2 | 50.8 | 1 |
| 50 | 400 | 300 | 0.74 | 0.56 | 0.99 | 49.5 | 50.5 | 1 |

| | | | | | | | | |
|---|---|---|---|---|---|---|---|---|
| 50 | 400 | 400 | 0.86 | 0.65 | 1.15 | 49.9 | 50.1 | 1 |
| 50 | 400 | 500 | 0.78 | 0.61 | 0.99 | 50.8 | 49.2 | 1 |
| 50 | 500 | 200 | 0.56 | 0.43 | 0.73 | 50.2 | 49.8 | 1 |
| 50 | 500 | 300 | 0.6 | 0.45 | 0.79 | 49.7 | 50.3 | 1 |
| 50 | 500 | 400 | 0.55 | 0.42 | 0.72 | 49.7 | 50.3 | 1 |
| 50 | 500 | 500 | 0.83 | 0.65 | 1.06 | 50.6 | 49.4 | 1 |
| 100 | 200 | 200 | 1.1 | 0.84 | 1.43 | 49 | 51 | 1 |
| 100 | 200 | 300 | 0.86 | 0.66 | 1.12 | 49.8 | 50.2 | 1 |
| 100 | 200 | 400 | 0.74 | 0.56 | 0.98 | 49.3 | 50.7 | 1 |
| 100 | 200 | 500 | 0.77 | 0.57 | 1.04 | 49.7 | 50.3 | 1 |
| 100 | 300 | 200 | 1.04 | 0.83 | 1.31 | 50.1 | 49.9 | 1 |
| 100 | 300 | 300 | 0.81 | 0.62 | 1.06 | 50.1 | 49.9 | 1 |
| 100 | 300 | 400 | 0.78 | 0.58 | 1.04 | 51 | 49 | 1 |
| 100 | 300 | 500 | 0.74 | 0.58 | 0.94 | 48.7 | 51.3 | 1 |
| 100 | 400 | 200 | 0.77 | 0.59 | 1.02 | 48.7 | 51.3 | 1 |
| 100 | 400 | 300 | 0.62 | 0.47 | 0.82 | 50.2 | 49.8 | 1 |
| 100 | 400 | 400 | 0.86 | 0.64 | 1.15 | 48 | 52 | 1 |
| 100 | 400 | 500 | 0.81 | 0.62 | 1.07 | 49.7 | 50.3 | 1 |
| 100 | 500 | 200 | 0.61 | 0.48 | 0.78 | 50.5 | 49.5 | 1 |
| 100 | 500 | 300 | 0.75 | 0.56 | 1.01 | 49.5 | 50.5 | 0 |
| 100 | 500 | 400 | 0.63 | 0.46 | 0.86 | 51.3 | 48.7 | 1 |
| 100 | 500 | 500 | 0.56 | 0.42 | 0.76 | 50.3 | 49.7 | 1 |

First column is the percentage of training sample size, second column is the batch size, third column is the number of training epochs, fourth column is the mean hazard ratio estimate of the outcome, fifth and sixth columns are lower and upper bounds of the 95% Confidence interval for the hazard ratio, seventh and eighth columns are the percentage of generated patients assigned intensive care and standard care arms, respectively, and in the final row, converged models based on graph inspection have value 1, non-converged, value 0. Rows in green indicate models with a hazard ratio and confidence intervals spanning 1, as seen in ACCORD. Abbreviations: CI: Confidence Interval, HR: Hazard Ratio, IC: Intensive Care, No.: Number, Pct. Percentage, SC: Standard Care

Table S9: Statistical Comparison of Non-Conditioned Twin Generation Approaches- First Aggregate Level

**A. All variables**

| Model | MAE | $Rank_{MAE}$ | $R^2$ | $Rank_{R^2}$ | RSME | $Rank_{RMSE}$ | SRMSE | $RANK_{SRMSE}$ | $\rho_{pearson}$ | $Rank_{\rho}$ | Rank Overall |
|---|---|---|---|---|---|---|---|---|---|---|---|
| **G. Copula** | **0.01** | **1** | **0.01** | **1** | -8.18 | 3 | **0.05** | **1** | **0.89** | **1** | **1.4** |
| **RCT-Twin-GAN** | 0.01 | 2 | 0.01 | 2 | -2.06 | 2 | 0.07 | 2 | 0.89 | 2 | 2 |
| **CTABGAN+** | 0.02 | 3 | 0.02 | 3 | **-1.87** | **1** | 0.14 | 3 | 0.87 | 5 | 3 |
| **CTGAN** | 0.04 | 4 | 0.05 | 4 | -27.74 | 4 | 0.19 | 5 | 0.77 | 6 | 4.6 |
| **CopulaGAN** | 0.04 | 5 | 0.05 | 5 | -7487.59 | 6 | 0.18 | 4 | 0.87 | 4 | 4.8 |
| **TVAE** | 0.05 | 6 | 0.06 | 6 | -92.10 | 5 | 0.19 | 6 | 0.88 | 3 | 5.2 |

**B. Continuous Variables**

| Model | MAE | $Rank_{MAE}$ | $R^2$ | $Rank_{R^2}$ | RSME | $Rank_{RMSE}$ | SRMSE | $RANK_{SRMSE}$ | $\rho_{pearson}$ | $Rank_{\rho}$ | Overall Rank |
|---|---|---|---|---|---|---|---|---|---|---|---|
| **G. Copula** | **0.01** | **1** | **0.01** | **1** | **0.96** | **1** | **0.1** | **1** | **0.98** | **1** | **1** |
| **RCT-Twin-GAN** | 0.01 | 2 | 0.02 | 2 | 0.95 | 2 | 0.2 | 2 | 0.98 | 2 | 2 |
| **CTABGAN+** | 0.03 | 6 | 0.04 | 6 | 0.58 | 6 | 0.4 | 6 | 0.92 | 6 | 6 |
| **CTGAN** | 0.03 | 5 | 0.04 | 5 | 0.82 | 5 | 0.4 | 5 | 0.92 | 5 | 5 |
| **CopulaGAN** | 0.02 | 3 | 0.03 | 4 | 0.84 | 4 | 0.3 | 4 | 0.94 | 4 | 3.8 |
| **TVAE** | 0.02 | 4 | 0.03 | 3 | 0.85 | 3 | 0.3 | 3 | 0.96 | 3 | 3.2 |

**C. Categorical Variables**

| Model | MAE | $Rank_{MAE}$ | $R^2$ | $Rank_{R^2}$ | RSME | $Rank_{RMSE}$ | SRMSE | $RANK_{SRMSE}$ | $\rho_{pearson}$ | $Rank_{\rho}$ | Overall Rank |
|---|---|---|---|---|---|---|---|---|---|---|---|
| **G. Copula** | **0.00** | **1** | **0.00** | **1** | -12.40 | 3 | **0.01** | **1** | 0.85 | 4 | **2** |
| **RCT-Twin-GAN** | 0.01 | 2 | 0.01 | 2 | -3.44 | 2 | 0.01 | 2 | 0.85 | 3 | 2.2 |
| **CTABGAN+** | 0.01 | 3 | 0.01 | 3 | **-3.00** | **1** | 0.03 | 3 | **0.85** | **1** | 2.2 |
| **CTGAN** | 0.05 | 4 | 0.05 | 4 | -40.92 | 4 | 0.10 | 4 | 0.69 | 6 | 4.4 |
| **CopulaGAN** | 0.06 | 5 | 0.06 | 5 | -10943.80 | 6 | 0.11 | 5 | 0.85 | 5 | 5.2 |
| **TVAE** | 0.07 | 6 | 0.07 | 6 | -135.00 | 5 | 0.13 | 6 | 0.85 | 2 | 5 |

Statistical Comparison of the marginal distribution of each variable of the non-conditioned digital twins generated by each method. All (A), continuous (B), or categorical (C) variables. Abbreviations: MAE: Mean Absolute Error, RSME: Root Mean Squared Error, SRMSE: Standardized Root Mean Squared Error, $\rho_{pearson}$**:** Pearson correlation, TVAE: Triplet-based Variational Autoencoder.

Table S10: Statistical Comparison of Non-Conditioned Twin Generation Approaches- Second Aggregate Level

**All variables**

| **Model** | **MAE** | **$Rank_{MAE}$** | **$R^2$** | **$Rank_{R^2}$** | **RSME** | **$Rank_{RMSE}$** | **SRMSE** | **$RANK_{SRMSE}$** | **$\rho_{pearson}$** | **$Rank_{\rho}$** | **Rank Overall** |
|---|---|---|---|---|---|---|---|---|---|---|---|
| **G. Copula** | **0.01** | **1** | **0.01** | **1** | **0.97** | **1** | **0.13** | **1** | **0.99** | **1** | **1** |
| **RCT-Twin-GAN** | 0.01 | 2 | 0.01 | 2 | 0.97 | 2 | 0.16 | 2 | 0.99 | 2 | 2 |
| **CTABGAN+** | 0.01 | 3 | 0.02 | 3 | 0.79 | 5 | 0.31 | 3 | 0.95 | 4 | 3.6 |
| **CTGAN** | 0.03 | 4 | 0.03 | 4 | 0.8 | 4 | 0.36 | 5 | 0.92 | 5 | 4.4 |
| **CopulaGAN** | 0.03 | 5 | 0.03 | 5 | 0.82 | 3 | 0.35 | 4 | 0.92 | 6 | 4.6 |
| **TVAE** | 0.04 | 6 | 0.05 | 6 | 0.71 | 6 | 0.43 | 6 | 0.96 | 3 | 5.4 |

Statistical Comparison of the marginal distribution of pairs of variables of the non-conditioned digital twins generated by each method. Abbreviations: MAE: Mean Absolute Error, RSME: Root Mean Squared Error, SRMSE: Standardized Root Mean Squared Error, $\rho_{pearson}$: Pearson correlation, TVAE: Triplet-based Variational Autoencoder.

Table S11: Comparison of Machine Learning Efficacy of Non-Conditioned Twin Generation Approaches

| | **Cat. V. Score** | **Cat. V. Rank** | **Cont. V. Score** | **Cont. V. Rank** | **Overall Rank** |
|---|---|---|---|---|---|
| **Original** | -0.514 | NA | 5.289 | NA | NA |
| **RCT-Twin-GAN** | 0.390 | 2 | **6.263** | **1** | **1.5** |
| **CTABGAN+** | **0.158** | **1** | 6.282 | 3 | 2 |
| **CopulaGAN** | 1.035 | 3 | 6.551 | 4 | 3.5 |
| **CTGAN** | 1.070 | 4 | 6.898 | 5 | 4.5 |
| **Gaussian Copula** | 1.351 | 5 | 6.263 | 2 | 3.5 |
| **TVAE** | 3.222 | 6 | 7.038 | 6 | 6 |

Comparison of Machine Learning Efficacy stratified by categorical or continuous variables. Abbreviations: Cat. V.: Categorical Variables, Cont. V.: Continuous Variables, TVAE: Triplet-based Variational Autoencoder.

Table S12: Baseline Characteristics of 5 SPRINT$_{EHR}$ Twins compared to original SPRINT treatment and placebo arms.

| Column | SPRINT C | SPRINT I | Twin 1 C | Twin 1 I | Twin 2 C | Twin 2 I | Twin 3 C | Twin 3 I | Twin 4 C | Twin 4 I | Twin 5 C | Twin 5 I |
|---|---|---|---|---|---|---|---|---|---|---|---|---|
| N | 4683 | 4678 | 1572 | 1558 | 1550 | 1580 | 1534 | 1596 | 1562 | 1568 | 1583 | 1547 |
| Age | 67[61,76] | 67[61,75] | 72[64,81] | 72[63,81] | 73[63,81] | 72[63,80] | 71[63,80] | 72[62,80] | 71[63,81] | 72[63,80] | 72[62,80] | 71[63,79] |
| Female | 1648(35) | 1684(35) | 781(49) | 772(49) | 767(49) | 812(51) | 749(48) | 792(49) | 760(48) | 728(46) | 785(49) | 749(48) |
| Black Race | 1423(30) | 1379(29) | 207(13) | 194(12) | 167(10) | 189(11) | 199(12) | 187(11) | 182(11) | 194(12) | 167(10) | 179(11) |
| Hispanic | 481(10) | 503(10) | 142(9) | 155(9) | 141(9) | 161(10) | 163(10) | 149(9) | 150(9) | 136(8) | 137(8) | 163(10) |
| White Race | 2701(57) | 2698(57) | 1186(75) | 1165(74) | 1199(77) | 1186(75) | 1131(73) | 1224(76) | 1194(76) | 1188(75) | 1230(77) | 1169(75) |
| BMI>30 | 1345(28) | 1402(29) | 456(29) | 437(28) | 444(28) | 474(30) | 428(27) | 429(26) | 437(27) | 458(29) | 445(28) | 416(26) |
| Systolic BP | 138[130,149] | 138[129,149] | 137[126,149] | 136[126,148] | 137[126,150] | 138[127,149] | 137[126,149] | 136[126,148] | 138[127,148] | 137[126,147] | 138[127,149] | 137[126,149] |
| Heart Rate | 65[58,74] | 65[58,73] | 74[64,87] | 74[64,87] | 73[63,87] | 73[64,87] | 74[64,87] | 74[65,86] | 73[64,86] | 74[64,86] | 74[64,87] | 74[64,86] |
| MI | 328(7) | 338(7) | 255(16) | 231(14) | 256(16) | 249(15) | 243(15) | 253(15) | 228(14) | 247(15) | 221(13) | 242(15) |
| LVH | 898(19) | 865(18) | 57(3) | 56(3) | 47(3) | 39(2) | 54(3) | 43(2) | 45(2) | 54(3) | 52(3) | 43(2) |
| On Statin | 2090(44) | 1990(42) | 1059(67) | 1011(64) | 1037(66) | 1071(67) | 978(63) | 1053(65) | 1038(66) | 1058(67) | 1046(66) | 1014(65) |
| GFR | 71[58,84] | 71[58,84] | 60[59,60] | 60[60,60] | 60[59,60] | 60[60,60] | 60[59,60] | 60[60,60] | 60[59,60] | 60[60,60] | 60[59,60] | 60[60,60] |
| LDL | 110[88,134] | 110[87,133] | 101[73,131] | 101[74,133] | 104[72,132] | 102[72,133] | 104[74,133] | 104[74,133] | 102[73,131] | 102[73,131] | 102[73,132] | 103[75,132] |
| Smoker | 602(12) | 640(13) | 178(11) | 150(9) | 139(8) | 159(10) | 152(9) | 177(11) | 159(10) | 153(9) | 162(10) | 154(9) |
| Family Hist. CVD | 2965(63) | 3063(65) | 393(25) | 406(26) | 419(27) | 394(24) | 384(25) | 424(26) | 401(25) | 409(26) | 406(25) | 366(23) |
| Angina | 588(12) | 625(13) | 292(18) | 272(17) | 283(18) | 284(17) | 289(18) | 274(17) | 261(16) | 276(17) | 275(17) | 290(18) |

First column includes covariates, second and third column includes original SPRINT population characteristics, and subsequent columns include SPRINT$_{EHR}$-Twins. Abbreviations: C: Control Arm, I: Intervention Arm, FH: Family History, CVD: Cardiovascular disease, BMI: Body Mass Index, GFR: Glomerular Filtration Rate, LVH: Left ventricular hypertrophy, LDL: low-density lipoprotein, MI: Myocardial infarction, SBP: Systolic Blood Pressure

Table S13: Baseline Characteristics of another 5 SPRINT$_{EHR}$ Twins compared to original SPRINT treatment and placebo arms.

| Column | SPRINT C | SPRINT I | Twin 1 C | Twin 1 I | Twin 2 C | Twin 2 I | Twin 3 C | Twin 3 I | Twin 4 C | Twin 4 I | Twin 5 C | Twin 5 I |
|---|---|---|---|---|---|---|---|---|---|---|---|---|
| N | 4683 | 4678 | 1568 | 1562 | 1591 | 1539 | 1556 | 1574 | 1543 | 1587 | 1558 | 1572 |
| Age | 67[61,76] | 67[61,75] | 72[63,80] | 71[62,80] | 72[63,81] | 73[63,80] | 73[64,80] | 72[63,80] | 71[63,80] | 71[62,80] | 72[63,80] | 72[63,80] |
| Female | 1648(35) | 1684(35) | 758(48) | 769(49) | 814(51) | 767(49) | 775(49) | 791(50) | 756(48) | 789(49) | 764(49) | 800(50) |
| Black Race | 1423(30) | 1379(29) | 183(11) | 193(12) | 175(10) | 183(11) | 187(12) | 173(10) | 198(12) | 188(11) | 181(11) | 183(11) |
| Hispanic | 481(10) | 503(10) | 163(10) | 162(10) | 159(9) | 137(8) | 147(9) | 137(8) | 146(9) | 161(10) | 158(10) | 138(8) |
| White Race | 2701(57) | 2698(57) | 1193(76) | 1170(74) | 1219(76) | 1171(76) | 1181(75) | 1214(77) | 1150(74) | 1197(75) | 1181(75) | 1213(77) |
| BMI>30 | 1345(28) | 1402(29) | 427(27) | 438(28) | 444(27) | 423(27) | 443(28) | 440(27) | 441(28) | 432(27) | 404(25) | 434(27) |
| Systolic BP | 138[130,149] | 138[129,149] | 137[126,148] | 137[127,148] | 137[126,149] | 137[126,148] | 137[127,150] | 138[126,149] | 138[127,148] | 137[126,148] | 138[127,149] | 137[127,147] |
| Heart Rate | 65[58,74] | 65[58,73] | 73[64,85] | 72[64,86] | 74[65,87] | 73[64,85] | 74[64,86] | 74[65,87] | 73[64,87] | 74[64,87] | 73[64,86] | 74[65,86] |
| MI | 328(7) | 338(7) | 223(14) | 259(16) | 252(15) | 245(15) | 221(14) | 246(15) | 223(14) | 234(14) | 252(16) | 251(15) |
| LVH | 898(19) | 865(18) | 34(2) | 46(2) | 47(2) | 51(3) | 42(2) | 51(3) | 44(2) | 48(3) | 43(2) | 23(1) |
| On Statin | 2090(44) | 1990(42) | 1059(67) | 1052(67) | 1049(65) | 1013(65) | 1033(66) | 1031(65) | 991(64) | 1040(65) | 1043(66) | 1012(64) |
| GFR | 71[58,84] | 71[58,84] | 60[59,60] | 60[60,60] | 60[60,60] | 60[59,60] | 60[59,60] | 60[59,60] | 60[60,60] | 60[60,60] | 60[60,60] | 60[59,60] |
| LDL | 110[88,134] | 110[87,133] | 100[72,129] | 104[73,131] | 104[75,133] | 103[71,134] | 103[74,131] | 103[75,132] | 104[76,136] | 103[75,134] | 102[71,132] | 102[74,132] |
| Smoker | 602(12) | 640(13) | 162(10) | 163(10) | 179(11) | 160(10) | 146(9) | 158(10) | 170(11) | 160(10) | 163(10) | 152(9) |
| Family Hist. CVD | 2965(63) | 3063(65) | 369(23) | 438(28) | 415(26) | 384(24) | 376(24) | 360(22) | 371(24) | 399(25) | 402(25) | 380(24) |
| Angina | 588(12) | 625(13) | 279(17) | 308(19) | 284(17) | 270(17) | 268(17) | 279(17) | 264(17) | 248(15) | 283(18) | 281(17) |

First column includes covariates, second and third column includes original SPRINT population characteristics, and subsequent columns include SPRINT$_{EHR}$-Twins. Abbreviations: C: Control Arm, I: Intervention Arm, FH: Family History, CVD: Cardiovascular disease, BMI: Body Mass Index, GFR: Glomerular Filtration Rate, LVH: Left ventricular hypertrophy, LDL: low-density lipoprotein, MI: Myocardial infarction, SBP: Systolic Blood Pressure

Table S14: Baseline Characteristics of 5 ACCORD$_{EHR}$ Twins compared to original ACCORD treatment and placebo arms.

| Column | ACCORD C | ACCORD I | Twin 1 C | Twin 1 I | Twin 2 C | Twin 2 I | Twin 3 C | Twin 3 I | Twin 4 C | Twin 4 I | Twin 5 C | Twin 5 I |
|---|---|---|---|---|---|---|---|---|---|---|---|---|
| N | 2371 | 2362 | 1347 | 1384 | 1329 | 1402 | 1390 | 1341 | 1366 | 1365 | 1352 | 1379 |
| Age | 62[57,67] | 61[57,67] | 66[58,73] | 66[59,73] | 67[60,74] | 66[59,73] | 66[59,73] | 66[59,73] | 67[59,73] | 67[59,73] | 66[59,73] | 67[60,74] |
| Female | 1130(47) | 1128(47) | 636(47) | 639(46) | 619(46) | 636(45) | 650(46) | 627(46) | 644(47) | 637(46) | 641(47) | 650(47) |
| Black Race | 580(24) | 547(23) | 275(20) | 287(20) | 245(18) | 265(18) | 285(20) | 257(19) | 252(18) | 272(19) | 292(21) | 263(19) |
| Hispanic | 170(7) | 160(6) | 91(6) | 111(8) | 97(7) | 96(6) | 102(7) | 94(7) | 116(8) | 104(7) | 105(7) | 82(5) |
| White Race | 1368(57) | 1413(59) | 887(65) | 905(65) | 907(68) | 938(66) | 928(66) | 882(65) | 933(68) | 903(66) | 894(66) | 958(69) |
| BMI>30 | 1213(51) | 1218(51) | 653(48) | 733(52) | 650(48) | 674(48) | 730(52) | 679(50) | 652(47) | 669(49) | 658(48) | 691(50) |
| Systolic BP | 138[130,149] | 138[128,149] | 139[126,149] | 137[125,147] | 138[125,150] | 138[126,148] | 138[126,149] | 138[126,148] | 138[126,150] | 138[127,149] | 138[126,149] | 138[127,148] |
| Heart Rate | 72[65,81] | 72[65,80] | 79[68,90] | 78[67,91] | 78[68,91] | 79[67,90] | 78[68,90] | 78[68,92] | 78[67,91] | 78[68,91] | 78[67,90] | 79[68,91] |
| MI | 341(14) | 309(13) | 216(16) | 220(15) | 214(16) | 225(16) | 223(16) | 227(16) | 245(17) | 228(16) | 214(15) | 213(15) |
| LVH | 121(5) | 113(4) | 41(3) | 40(2) | 34(2) | 42(2) | 51(3) | 44(3) | 30(2) | 39(2) | 44(3) | 40(2) |
| On Statin | 1562(65) | 1511(63) | 1098(81) | 1138(82) | 1094(82) | 1126(80) | 1143(82) | 1098(81) | 1127(82) | 1147(84) | 1114(82) | 1131(82) |
| GFR | 89[75,105] | 89[75,105] | 60[59,60] | 60[59,60] | 60[58,60] | 60[59,60] | 60[60,60] | 60[58,60] | 60[57,60] | 60[59,60] | 60[59,60] | 60[58,60] |
| LDL | 105[85,129] | 106[86,133] | 96[71,121] | 93[70,122] | 96[71,124] | 96[71,123] | 95[70,123] | 96[71,122] | 96[70,123] | 96[71,125] | 95[70,124] | 96[72,124] |
| Smoker | 269(11) | 279(11) | 164(12) | 148(10) | 144(10) | 170(12) | 168(12) | 167(12) | 148(10) | 142(10) | 161(11) | 145(10) |
| Family Hist. CVD | 743(31) | 761(32) | 366(27) | 387(27) | 359(27) | 377(26) | 392(28) | 354(26) | 374(27) | 343(25) | 395(29) | 392(28) |
| Angina | 266(11) | 270(11) | 181(13) | 238(17) | 196(14) | 169(12) | 235(16) | 192(14) | 191(13) | 199(14) | 198(14) | 184(13) |

First column includes covariates, second and third column includes original ACCORD population characteristics, and subsequent columns include ACCORD$_{EHR}$-Twins. Abbreviations: C: Control Arm, I: Intervention Arm, FH: Family History, CVD: Cardiovascular disease, BMI: Body Mass Index, GFR: Glomerular Filtration Rate, LVH: Left ventricular hypertrophy, LDL: low-density lipoprotein, MI: Myocardial infarction, SBP: Systolic Blood Pressure

Table S15: Baseline Characteristics of another 5 ACCORD$_{EHR}$ Twins compared to original ACCORD treatment and placebo arms.

| Column | ACCORD C | ACCORD I | Twin 1 C | Twin 1 I | Twin 2 C | Twin 2 I | Twin 3 C | Twin 3 I | Twin 4 C | Twin 4 I | Twin 5 C | Twin 5 I |
|---|---|---|---|---|---|---|---|---|---|---|---|---|
| N | 2371 | 2362 | 1353 | 1378 | 1341 | 1390 | 1332 | 1399 | 1340 | 1391 | 1303 | 1428 |
| Age | 62[57,67] | 61[57,67] | 67[59,73] | 66[59,73] | 67[59,74] | 66[59,73] | 66[58,72] | 66[58,73] | 66[59,73] | 66[59,73] | 66[59,74] | 67[59,73] |
| Female | 1130(47) | 1128(47) | 639(47) | 604(43) | 648(48) | 651(46) | 630(47) | 652(46) | 601(44) | 651(46) | 590(45) | 646(45) |
| Black Race | 580(24) | 547(23) | 270(19) | 299(21) | 276(20) | 283(20) | 271(20) | 270(19) | 278(20) | 271(19) | 256(19) | 296(20) |
| Hispanic | 170(7) | 160(6) | 111(8) | 112(8) | 96(7) | 108(7) | 101(7) | 118(8) | 115(8) | 91(6) | 109(8) | 93(6) |
| White Race | 1368(57) | 1413(59) | 890(65) | 878(63) | 873(65) | 904(65) | 877(65) | 918(65) | 889(66) | 926(66) | 849(65) | 954(66) |
| BMI>30 | 1213(51) | 1218(51) | 663(49) | 685(49) | 671(50) | 696(50) | 674(50) | 661(47) | 628(46) | 687(49) | 656(50) | 709(49) |
| Systolic BP | 138[130,149] | 138[128,149] | 138[127,149] | 139[127,149] | 138[125,148] | 137[125,148] | 137[125,149] | 138[127,149] | 138[126,149] | 137[126,147] | 138[126,148] | 138[126,148] |
| Heart Rate | 72[65,81] | 72[65,80] | 78[67,91] | 78[68,90] | 79[68,91] | 78[68,90] | 78[69,91] | 78[68,91] | 78[68,91] | 78[68,90] | 78[68,90] | 79[68,92] |
| MI | 341(14) | 309(13) | 241(17) | 228(16) | 211(15) | 224(16) | 211(15) | 221(15) | 206(15) | 226(16) | 215(16) | 242(16) |
| LVH | 121(5) | 113(4) | 37(2) | 39(2) | 41(3) | 40(2) | 40(3) | 38(2) | 39(2) | 42(3) | 33(2) | 42(2) |
| On Statin | 1562(65) | 1511(63) | 1127(83) | 1143(82) | 1075(80) | 1127(81) | 1087(81) | 1129(80) | 1090(81) | 1134(81) | 1080(82) | 1182(82) |
| GFR | 89[75,105] | 89[75,105] | 60[59,60] | 60[59,60] | 60[59,60] | 60[59,60] | 60[59,60] | 60[59,60] | 60[57,60] | 60[59,60] | 60[59,60] | 60[59,60] |
| LD | 105[85,129] | 106[86,133] | 96[71,124] | 96[70,121] | 96[70,123] | 93[70,122] | 96[71,124] | 95[70,123] | 96[71,123] | 98[72,124] | 94[69,121] | 99[70,126] |
| Smoker | 269(11) | 279(11) | 156(11) | 164(11) | 142(10) | 161(11) | 160(12) | 159(11) | 160(11) | 161(11) | 157(12) | 171(11) |
| Family Hist. CVD | 743(31) | 761(32) | 383(28) | 364(26) | 336(25) | 384(27) | 362(27) | 380(27) | 344(25) | 403(28) | 359(27) | 385(26) |
| Angina | 266(11) | 270(11) | 191(14) | 187(13) | 185(13) | 210(15) | 205(15) | 217(15) | 208(15) | 204(14) | 170(13) | 207(14) |

First column includes covariates, second and third column includes original ACCORD population characteristics, and subsequent columns include ACCORD$_{EHR}$-Twins. Abbreviations: C: Control Arm, I: Intervention Arm, FH: Family History, CVD: Cardiovascular disease, BMI: Body Mass Index, GFR: Glomerular Filtration Rate, LVH: Left ventricular hypertrophy, LDL: low-density lipoprotein, MI: Myocardial infarction, SBP: Systolic Blood Pressure

Table S16: ICD-10-CDM codes used to extract EHR cohorts and covariates.

| Covariate | Variables | OMOP Table Used and Variable Name |
|---|---|---|
| Age | | Person, date of birth and death datettime |
| Black Race | ‘Black or African American’ | Person, race source value |
| Body Mass Index | 'BMI (Calculated)','BMI (Calculated)','[Retired] BMI (kg/m2)' | Measurement, measurement source value |
| Family History Cardiovascular Disease | 4148407,4170147 | Condition, condition concept id |
| Female | ‘Female’ | Person, gender source value |
| GFR | 'eGFR (NON African-American)','eGFR (Afr Amer)','eGFR (AFRICAN-AMERICAN)','EGFR','eGFR','eGFR (Creatinine)','POC eGFR (Afr Amer)','POC eGFR (NON African-American)','POC eGFR (Creatinine)','POC eGFR' | Measurement, measurement source value |
| Heart Rate | 'ECG - HEART RATE','Heart Rate' | Measurement, measurement source value |
| Hypertension | 320128,44782429,312648,317898,44782690,319826,4028741,4058286,317895,4289933,44783644,4167493,4118910,321074,36713024,4311246,4291933,4302591,4016922,36712757,141084,4283352,4269358,44782689,37016726,46273804,44782691,45757788,4159755,4017170,37018886,4209293,4227607,4110947,4167358,318437,4249016,4217486,314958,135601,4049389,136743,321638,4057979,46273636,44783643,4263067,4058987,46270353,4062811,4062550,4253928,4179379,4276511,4180283,4034031,4034094,46273514,4221991,46270355,44782692,4146627,4218088,4032952,4242878,42538697,4269035,314423,4277110,4088880,4110948,42538946,4178312,4024560,37311148,42873163,321080,4108213,4262182,4279525,4006325,37311147,45772751,45771064,45757445,4322735,45757446,45757447,4219323,4061667 | Condition, condition concept id |
| Left Ventricular hypertrophy | 4184746,4322893,4102533,3654813,3654814,3654815 | Condition, condition concept id |
| Myocardial Infarction | 4329847,37309626,4170094,4147400,314666,4270024,312327,4296653,4163874,43020460,46270163,434376,438170,44784441,4108680,45766241,37311078,46270162,438438,37395582,438447,441579,436706,45766114,45773170,4108218,439693,4200113,4121467,4119949,4108677,45766113,45766116,44782769,4124686,4176766,3654465,3661520,765132,438172,45766075,4108679,4064348,4322145,43530755,46270159,3661504,4108220,4108678,37016102,319039,46274044,46270161,3661502,4108219,4267568,45766151,37109912,4119950,44782712,4108217,4243372,35611570,35611571,4126801,367129854051874,37109911,46270160,45771322,46270158,4178129 | Condition, condition concept id |
| Statin | 'Antihyperlipidemic - HMG CoA Reductase Inhibitors (statins)', 'Antihyperlipidemic - HMG CoA Reductase Inhibitor and Niacin Comb', 'Antihyperlipidemic-HMG CoA Reduct Inhib and Cholesterol Absorp Inhibit', 'Antihyperlipidemic HMG CoA Reduct Inhib and Calcium Channel Blocker' | Drug, drug class source value |
| Type 2 Diabetes Mellitus | 4193704,201826,4200875,40485020,4099651,4304377,4230254,45757508,37016349,443732,43531578,376065,4130162,443729,37017432,37016354,443731,45757363,443733,45757435,43530690,4226121,43531616,37016768,45770830,43530685,4222876,4577088 | Condition, condition concept id |

| | |
|---|---|
| | 1,35626070,4222415,36712687,36712686,43531563,36714116,201530,43530656,45757449,43531653,43531577,4215719,4196141,37312205,43530689,45769905,45773064,43531010,4140466,4228443,43531562,43531651,43531566,4129519,37018728,45757277,45757499,45770880,443734,45757278,45769836,45769888,45769906,4063043,43531588,4177050,36712670,37309630,43531559,43531564,45763582,45757445,45757280,45771064,43531608,45757446,45769875,45769828,45769872,45770831,45757444,37018912,4321756,45757450,45769890,45757447,4142579,45772060,45757392,36717156,4223739,45757075] |

Table S17: Covariates and bounds for continuous variables.

| | Data Encoding | | | |
|---|---|---|---|---|
| Covariate | Type | Bounds (Original) | Bounds (Winsorized) | Discrete |
| Age | Continuous | [0, 90] | [50, 90] | True |
| GFR | Continuous | [0, 200] | [30, 115] | False |
| Heart Rate | Continuous | [20, 150] | [45, 95] | True |
| LDL Cholesterol | Continuous | [0, 360] | [55, 190] | True |
| Systolic BP | Continuous | [60, 240] | [110, 175] | True |
| Time to Outcome | Continuous | [0, 5] | [0, 5] | False |
| Angina | Categorical | | - | - |
| Black Race | Categorical | | - | - |
| BMI | Categorical | | - | - |
| Current Smoker | Categorical | | - | - |
| Family History | Categorical | | - | - |
| Female Sex | Categorical | | - | - |
| Group | Categorical | | - | - |
| Hispanic Race | Categorical | | - | - |
| LVH | Categorical | | - | - |
| MI | Categorical | | - | - |
| Outcome | Categorical | | - | - |
| Statin | Categorical | | - | - |
| White Race | Categorical | | - | - |

Fourth column shows bounds winsorized at 2.5% and 97.5% percentiles. Abbreviations: T: Treatment Arm, P: Placebo Arm, FH: Family History, CVD: Cardiovascular disease, BMI: Body Mass Index, GFR: Glomerular Filtration Rate, LVH: Left ventricular hypertrophy, LDL: low-density lipoprotein, MI: Myocardial infarction, SBP: Systolic Blood Pressure

Table S18: Summary of baseline features available from both SPRINT and ACCORD.

| Covariate | Chosen for Model? (Y/N) | Reason For Inclusion/Exclusion |
|---|---|---|
| **Demographics** | | |
| Age | **Y** | Cardiac Risk Factor, Available in EHR |
| Female Sex | **Y** | Cardiac Risk Factor, Available in EHR |
| Race (White, Black, Hispanic, Other) | **Y** | Cardiac Risk Factor, Available in EHR |
| **Conditions** | | |
| Left ventricular hypertrophy (LVH) by electrocardiography | **Y** | Cardiac Risk Factor and an EKG value, Available in EHR |
| Obesity (Body mass index (BMI) greater than 30) | **Y** | Cardiac Risk Factor, Available in EHR |
| Carotid disease | **N** | Overlapping risk factor with cholesterol level |
| Cardiovascular Disease (CVD) | **N** | Overlapping risk factor with angina, FH of CVD, MI |
| Diabetes | **N** | Exclusion criterium in SPRINT |
| Known 50% or greater coronary, carotid, or peripheral stenosis | **N** | Overlapping risk factor with cholesterol level |
| History of myocardial infarction (MI) | **N** | Cardiac Risk Factor, Available in EHR |
| Stroke | **N** | Overlapping risk factor with current smoker, previous MI |
| **Medications** | | |
| Aspirin Use | **N** | Overlapping with smoking, FH of CVD, previous MI |
| Statin Use | **Y** | Cardiac medication, Available in EHR |
| **Family History** | | |
| Family History of Cardiovascular Disease (FH of CVD) | **Y** | Cardiac Risk Factor, Available in EHR |
| **Symptoms** | | |
| Angina | **Y** | Only variable in symptom category, Available in EHR |
| **Social History** | | |
| Smoking | **Y** | Only variable in social history category, Available in EHR |
| **Procedures** | | |
| History of coronary revascularization | **N** | Overlapping risk factor with previous MI |
| **Vital Signs** | | |

| | | |
|---|---|---|
| Systolic blood pressure | **Y** | Cardiac risk factor, affected by intervention, available in EHR |
| Diastolic blood pressure | **N** | Other vital signs used |
| Heart rate | **Y** | Associated with |
| **Laboratory and EKG Values** | | |
| Creatinine (serum) | **N** | Overlapping with eGFR |
| Total cholesterol (serum) | **N** | Overlapping with LDL cholesterol |
| Fasting glucose (serum) | **N** | Limited distribution overlap between SPRINT and ACCORD cohorts |
| Glomerular filtration rate (eGFR) (serum) | **Y** | Proxy for kidney disease, cardiac risk factor |
| High-density lipoprotein (serum) | **N** | Overlapping with LDL cholesterol |
| Potassium (serum) | **N** | Overlapping with eGFR |
| Low-density lipoprotein (serum) | **N** | Overlapping with LDL cholesterol |
| Sodium (serum) | **N** | eGFR more relevant to intervention |
| Triglycerides (serum) | **N** | Overlapping with LDL cholesterol |
| Urine microalbumin/creatinine ratio | **N** | Overlapping with eGFR |
| Cornell voltage | **N** | Overlapping with LVH |
| QRS duration | **N** | LVH is representative EKG value more related to intervention |

Table S19: Pairs in Directed Acyclic Graph.

| Arrow From | Arrow To |
|---|---|
| Age | Outcome |
| Age | Time to outcome |
| Age | Myocardial Infarction |
| Age | Glomerular Filtration Rate |
| Age | Current Smoker |
| Age | Statin Use |
| Age | LDL Cholesterol |
| Age | Heart Rate |
| Angina | Outcome |
| Angina | Time to outcome |
| Angina | Myocardial Infarction |
| Angina | Statin Use |
| Black Race | Outcome |
| Black Race | Body Mass Index |
| Black Race | Systolic Blood Pressure |
| Black Race | Hispanic Ethnicity |
| Black Race | Left Ventricular Hypertrophy |
| Black Race | Glomerular Filtration Rate |
| Body Mass Index | Outcome |
| Body Mass Index | Time to outcome |
| Body Mass Index | Systolic Blood Pressure |
| Body Mass Index | Age |
| Current smoker | Outcome |
| Current smoker | Time to outcome |
| Current Smoker | Angina |
| Current Smoker | Myocardial Infarction |
| Family History of Cardiovascular Disease | Outcome |
| Family History of Cardiovascular Disease | Time to outcome |
| Family History of Cardiovascular Disease | Myocardial Infarction |
| Female | Outcome |
| Female | Time to outcome |
| Female Sex | Myocardial Infarction |
| Female Sex | Current Smoker |
| Female Sex | LDL Cholesterol |
| Glomerular Filtration Rate | Outcome |
| Glomerular Filtration Rate | Time to outcome |
| Group | Outcome |
| Group | Time to outcome |
| Heart Rate | Outcome |
| Heart Rate | Time to outcome |
| Hispanic ethnicity | Outcome |
| Hispanic ethnicity | Time to outcome |

| | |
|---|---|
| Hispanic Ethnicity | Body Mass Index |
| Hispanic Ethnicity | Systolic Blood Pressure |
| Hispanic Ethnicity | Glomerular Filtration Rate |
| LDL Cholesterol | Outcome |
| LDL Cholesterol | Tme to outcome |
| LDL Cholesterol | Myocardial Infarction |
| LDL Cholesterol | Statin use |
| LDL Cholesterol | Angina |
| Left Ventricular Hypertrophy | Outcome |
| Left Ventricular Hypertrophy | Time to outcome |
| Myocardial Infarction | Outcome |
| Myocardial Infarction | Time to outcome |
| Myocardial Infarction | Statin use |
| Outcome | Time to outcome |
| Statin use | Outcome |
| Statin use | Time to outcome |
| Systolic Blood Pressure | Outcome |
| Systolic Blood Pressure | Time to outcome |
| Systolic Blood Pressure | Left Ventricular Hypertrophy |
| White Race | Outcome |
| White Race | Body Mass Index |
| White Race | Systolic Blood Pressure |
| White Race | Black Race |
| White Race | Hispanic Ethnicity |
| White Race | Glomerular Filtration Rate |

Left column represents the source node, and right column represents the receiver node

Table S20: RCT-Twin-GAN Parameter search grid

| Hyperparameter Grid Search | |
|---|---|
| Hyperparameters | Search Space |
| Batch Size | 400, 700, 1000, 1300 |
| Epochs | 300, 400, 500 |
